# Comparing and combining TSPO-PET tracers in tauopathies

**DOI:** 10.1101/2025.07.23.25332044

**Authors:** Harry Crook, Nicolai Franzmeier, Nesrine Rahmouni, Johannes S Gnörich, Tim Fryer, Young Hong, Sebastian N Roemer-Cassiano, Carla Palleis, Alexandra Strauss, P Simon Jones, Franklin I. Aigbirhio, Robert Hopewell, Boris-Stephan Rauchmann, Gassan Massarweh, Robert Perneczky, Johannes Levin, Günter U Höglinger, James B Rowe, John T O’Brien, Pedro Rosa-Neto, Matthias Brendel, Maura Malpetti

## Abstract

**Purpose:** Neuroinflammation is a key pathological driver in neurodegenerative diseases, including Alzheimer’s disease (AD) and Progressive Supranuclear Palsy (PSP). Positron emission tomography (PET) with tracers targeting the translocator protein (TSPO) enables the in vivo quantification of microgliosis. Different TSPO tracers have shown similar patterns across disease-specific cohorts. However, direct quantitative comparisons between commonly used TSPO-PET tracers in tauopathies have not been performed. Here, we apply our TSPO-PET standardization pipeline across clinically matched AD and PSP cohorts, to quantify, compare and combine multi-centre TSPO-PET data.

**Methods:** Patients with PSP were scanned with either [^11^C]PK11195 or [^18^F]GE-180 at one of two centres, while patients with AD and control participants were scanned with either [^11^C]PK11195, [^18^F]GE-180 or [^11^C]PBR28 at one of three centres. A standardised pre-processing pipeline was implemented and participant standardised uptake volume ratio (SUVR) values were z-scored using tracer-specific control participant values. In a data-driven approach, dissimilarity analyses were employed to assess differences between tracers across clinically matched cohorts.

**Results:** In PSP, dissimilarity analysis suggested that [^11^C]PK11195 and [^18^F]GE-180 were comparable following standardisation. In AD, comparability across tracers was less robust, with [^11^C]PK11195 and [^18^F]GE-180 being most comparable, followed by [^18^F]GE-180 vs [^11^C]PBR28, then by [^11^C]PK11195 vs [^11^C]PBR28.

**Conclusion:** The pipeline was effective at harmonising TSPO-PET tracers and standardising the regional quantification of neuroinflammation in clinically matched cohorts of PSP, while the standardisation pipeline results were less robust across AD cohorts.

## Introduction

The 18kDa translocator protein (TSPO) is expressed on the outer mitochondrial membrane within cells of organs including the kidneys, lungs, heart, and brain [1]. While the exact role of TSPO in the central nervous system remains elusive, its upregulation is linked to the proliferation, migration, and phagocytic functions of microglia, as well as the maintenance of mitochondrial homeostasis and the release of inflammatory cytokines [2]. In neurodegenerative disease, TSPO has been shown to be upregulated in microglia [3–5], and to a lesser extent in astrocytes and vasculature. For these reasons, TSPO has been considered a marker of neuroinflammation. Hence, positron emission tomography (PET) radiotracers have been developed that specifically target TSPO to measure *in vivo* tissue neuroinflammatory response in patients with dementia and other conditions.

Neuroinflammation is a key pathological feature of all neurodegenerative diseases [6], and TSPO-PET has been shown to be a useful marker to quantify neuroinflammation *in vivo* across neurodegenerative diseases. Indeed, heightened levels of neuroinflammation, as measured by TSPO-PET binding, has been associated with a greater rate of cognitive decline and disease progression in progressive supranuclear palsy (PSP) [7], frontotemporal dementia [8], Alzheimer’s disease (AD) dementia, and mild cognitive impairment (MCI) [9, 10], while high TSPO-PET binding in early stages increases the risk of developing dementia in patients with Parkinson’s disease [11]. Moreover, neuroinflammation has been associated with brain network dysfunction, as measured with resting state functional magnetic resonance imaging (MRI) [12], and the neuroinflammatory signal strongly correlates with neuropathological substrates topographically, especially tau protein, in both primary and secondary tauopathies [13–18]. Across diseases, patients with high TSPO-PET binding also exhibit elevated levels of inflammatory proteins in cerebrospinal fluid [19] and serum [20].

Several tracers have been developed, including the carbon-11 radiolabelled first generation tracers such as PK11195 (1-(2-chlorophenyl)-N-methyl-N-(1-methylpropyl)-3-isoquinoline carboxamide) and Ro5-4864 (4′-chlorodiazepam). While [^11^C]Ro5-4864 binding varies due to temperature and species, [^11^C]PK11195 has a high affinity and selectivity to TSPO and has been widely used to study neuroinflammation [21]. Despite the success of [^11^C]PK11195, its drawbacks, including a short half-life, low signal-to-noise ratio, and variable kinetics, have encouraged the development of second- and third-generation tracers. These tracers include [^11^C]PBR28, [^11^C]ER176, as well as the fluorine-18 radiolabelled tracers [^18^F]DPA-714, and [^18^F]GE-180, which have extended half-lives compared to carbon-11 labelled tracers. Although the second- and third-generation tracers display greater signal-to-noise than their predecessors, a common single nucleotide polymorphism (rs6971) in the TSPO gene affects their binding affinity, meaning that participants can have a high-, mixed-, or low-affinity binding and those in the latter group cannot be effectively imaged with these more recently developed tracers [22–25]. Despite its lower signal-to-noise ratio, [^11^C]PK11195 is minimally affected by this genetic variable.

Despite differences across tracers, previous studies in clinically matched cohorts of patients with tauopathies have identified regional patterns of increased TSPO-PET signal, which parallel known regional patterns of tau aggregates. For example, when compared to healthy volunteers, temporal, parietal, and occipital regions demonstrate elevated TSPO-PET binding in AD [26–28], while in the primary tauopathy PSP, the thalamus, putamen, pallidum, and midbrain are the most affected regions [15, 26, 29].

Although head-to-head studies comparing different TSPO-PET tracers have been conducted in small groups of healthy controls [30, 31], direct and quantitative comparisons from distinct patient cohorts with tauopathies imaged using various TSPO-PET tracers have not been reported. The goal of this study, therefore, is to enhance the utility of TSPO-PET imaging in tauopathies by harmonising the processing pipelines in order to compare and combine tracers. Should this methodology prove effective, it will allow for the creation of multicentre databases larger than those achievable with institution-specific cohorts, as well as a harmonised pipeline and universal scales to estimate microglial activation severity irrespective of the TSPO-PET tracer in use. Here, we describe the harmonised processing pipeline and the dissimilarity analyses that we have undertaken to assess the strength and validity of this pipeline. We have included patients with primary tauopathy PSP and secondary tauopathy AD to test this process across three centres and TSPO-PET tracers.

## Material and methods

### Participant cohorts

Patients with a clinical diagnosis of PSP Richardson’s syndrome (PSP-RS) [32], or of MCI due to AD (MCI-AD) and/or Alzheimer’s Dementia [33, 34], alongside age- and sex-matched cognitively unimpaired controls, were recruited across three centres: the University of Cambridge and Cambridge University Hospitals NHS Foundation Trust, Cambridge, UK; LMU Hospital, Ludwig-Maximilians Universität München, Munich, Germany; and McGill University and Montreal Neurological Institute Hospital, Montreal, Canada. All participants provided written informed consent according to the Declaration of Helsinki.

Specifically, 18 patients with PSP-RS, 32 patients with AD (14 AD and 18 MCI-AD), and 15 control participants underwent [^11^C]PK11195 PET in Cambridge; 17 patients with PSP-RS, 40 patients with AD, and 19 control participants underwent [^18^F]GE-180 PET in Munich; 25 patients with AD (11 AD and 14 MCI-AD) and 25 control participants underwent [^11^C]PBR28 PET in Montreal.

All participants undertook a detailed clinical and cognitive assessment. All patients with PSP-RS completed the PSP rating scale [35], while patients with AD completed either the Mini-mental state examination (MMSE) [36] or the Montreal cognitive assessment (MoCA) [37].

### PET acquisition

[^11^C]PK11195 PET was undertaken at the Wolfson Brain Imaging Centre at the University of Cambridge, United Kingdom. Scans were performed on GE Advance and GE Discovery 690 PET/CT (GE Healthcare) scanners with a 15-min 68Ge/68Ga transmission computed tomography (CT) scan for attenuation correction. 500 MBq of [^11^C]PK11195 was injected intravenously over 30-seconds at the onset of a 75-minute dynamic scan.

[^18^F]GE-180 PET was undertaken at Ludwig Maximilian University of Munich, Germany. These scans were completed on a Siemens Biograph True point 64 PET/CT or a Siemens mCT PET/CT scanner (Siemens, Erlangen, Germany) with CT used in combination for attenuation correction. 185 MBq of [^18^F]GE-180 was injected intravenously and PET images acquired statically at 60-80 minutes post-injection.

[^11^C]PBR28 PET was undertaken at the Brain Imaging Centre, Montreal Neurological Institute, McGill University, Canada. Scans were performed on a Siemens high-resolution research tomograph with a 6-min transmission scan performed at the end of each PET emission acquisition for attenuation correction. A mean dose of 384 MBq of [^11^C]PBR28 was injected intravenously, and emission PET images were acquired from 0-90 minutes and reconstructed from 60-90 minutes.

All TSPO-PET data was visually checked for scan quality and artifacts prior to preprocessing. All participants that were low affinity binders due to the TSPO polymorphism rs6971 and underwent [^18^F]GE-180 PET or [^11^C]PBR28 PET were removed from the analysis [24].

### Harmonised PET pre-processing pipeline

A 40-70 minute time window after injection from the dynamic [^11^C]PK11195 PET scans was considered to calculate the standardised uptake value ratio (SUVR), which was selected based on the correlation between SUVR and distribution volume ratio for various SUVR time windows, as well as the stability of the SUVR values. The [^18^F]GE-180 signal, meanwhile, was recorded 60-80 minutes after injection and [^11^C]PBR28 was recorded at 60-90 minutes post-injection to calculate SUVR.

Each participants [^11^C]PK11195 PET image was co-registered to their corresponding T1-weighted MR image and kept in subject space, while each [^18^F]GE-180 and [^11^C]PBR28 PET images were transformed to Montreal Neurological Institute (MNI) space. Inverse transform parameters were applied to a modified version of the Hammers Atlas [38] to bring the regions of interest (ROI) to scan-specific space. The modified version of the atlas allowed for the inclusion of brainstem regions into the analysis which are important regions to include for the assessment of patients with PSP-RS.

All PET images were intensity normalised to mean tracer uptake of a shared reference region of the cerebellar grey matter in order to determine the SUVR. The reference region was defined from the Hammers cerebellum VOI with manual adjustment, excluding the dentate nucleus, and superior and posterior layers. In order to maintain harmony between the processing of the tracers, vascular binding correction and partial volume correction was not applied. Averaged SUVR values across grey and white matter from the Hammers atlas ROIs were extracted from each participant.

### Z-scoring

Corresponding left and right regional SUVR values from the Hammers atlas were averaged to obtain bilateralised regional SUVR values. To standardise the SUVR scales, z-scores for each region were calculated for each patient, based on centre-specific control participants. The following formula was employed to calculate Z-scores:

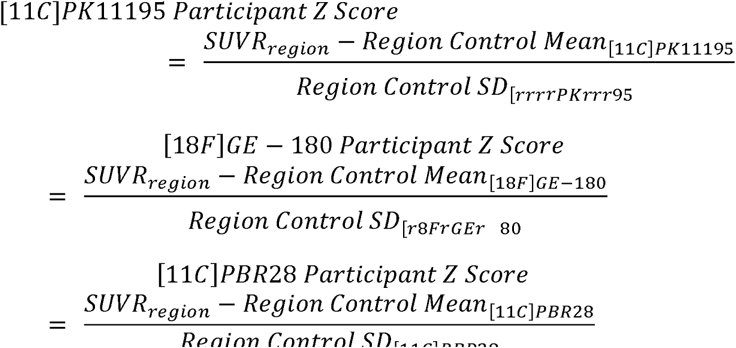

### Statistical analysis

Due to some regional z-scored SUVR values having a non-normal distribution and not meeting assumptions for parametric tests, non-parametric statistical tests were employed. We applied the same combination of dissimilarity analyses separately for PSP-RS and AD cohorts, to determine whether any tracer-specific differences could be identified within disease groups.

First, to compare [^11^C]PK11195 and [^18^F]GE-180 regional z-scores in the PSP-RS and control groups two-sided, Mann-Whitney U tests were employed, while Kruskal-Wallis and post-hoc Dunn’s tests were used to compare regional z-scores of all three tracers in the AD and control groups. These analyses were adjusted for multiple comparisons using false discovery rate (FDR) correction.

Second, a full factorial analysis was implemented to systematically examine the effects of the diagnostic group, TSPO-PET tracer, and brain region, on the regional z-score. A main effects model (z-score ∼ diagnostic group + tracer + region + age + sex), as well as a model to examine two-factor interactions (z-score ∼ diagnostic group:tracer + region + age + sex) and three-factor interactions (z-score ∼ diagnostic group:tracer:region + age + sex) was applied. For the two- and three-factor interaction models, family-wise error corrected post-hoc estimated marginal means analyses were employed to identify interaction effects. Due to having z-scores closest to 0 for both PSP-RS and AD groups, the precentral gyrus was used as the reference region for this model to compare to all other brain regions.

Then, Euclidean distance was used to calculate the distance between each participants z-scores in forty-one-dimensional space, based on 41 brain regions of interest, and clustering algorithms were run to visualise the distance and any clustering of participants based on tracer. Hierarchical agglomerative clustering was used as a data exploration tool to assess how closely participants cluster, with a dendrogram fitted to visualise any clustering based on PET tracer. K-means clustering of Euclidean distances was also applied with two clusters set for the PSP-RS group and three for the AD group.

Finally, to observe the pairwise dissimilarities of regional z-scores between tracers, representational similarity analysis was implemented using correlation distance (1 minus the correlation coefficient) for each brain region. To further explore the patterns in the dissimilarity matrices computed by the representational similarity analysis, pattern similarity using Spearman’s correlation between tracer matrices was performed to evaluate the correlation between dissimilarities observed in the matrices. Lastly, permutation testing with 5000 simulations was implemented to assess the likelihood that any of the observed correlations between the tracers could have occurred by chance.

All statistical analyses were conducted in R version 4.3.2 (2023-10-31) [39]. A significance level of p<0.05 was applied throughout.

## Results

### Participants and demographics

Participant numbers and demographics are listed in table 1. For 8 out of 40 patients with AD who underwent [^18^F]GE-180 PET, MMSE scores were obtained with score conversion from MoCA using a pre-established approach [40].

### Harmonisation across PSP-RS cohorts: [^11^C]PK11195 and [^18^F]GE-180 results

Several dissimilarity analyses were run to determine whether any tracer-specific differences could be identified.

### Pairwise comparisons

No differences in z-scores were observed between tracers for any brain region in control participants. In PSP-RS, there were no differences in z-scores between tracers for any region except the subcallosal area (p=0.04) (Figure 1).

**Fig. 1:**
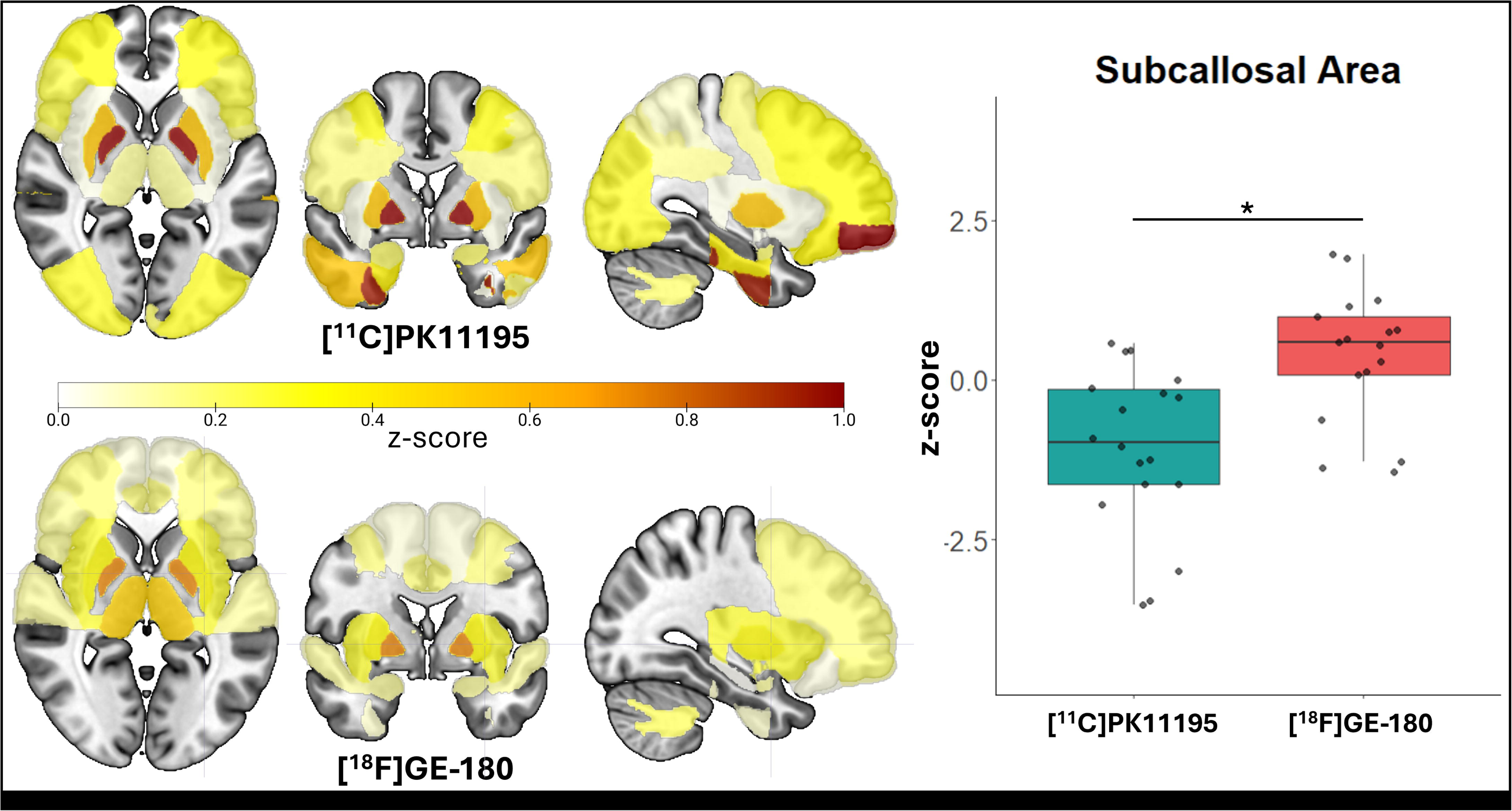
z-score brain plots of patients with PSP-RS. In patients with PSP-RS, heightened TSPO-PET z-scores were observed across the brain compared to controls. Differences between the two tracers was seen in the subcallosal area only. *=p<0.05

### Full factorial analysis

The main effects model (z-score ∼ diagnostic group + tracer + region + age + sex) was significant (F(44, 2784)=3.881, p<0.001). There was a significant main effect of tracer on predicting z-score ([^11^C]PK11195 - [^18^F]GE-180, est.=0.089, p=0.041), suggesting a difference between the two tracers. Main effects of diagnostic group (est.=0.136, p=0.002), age (est.=0.033, p<0.001), and sex (est.=- 0.245, p<0.001) to predict z-score were also observed. Of the 40 brain regions considered, the anterior superior temporal gyrus (est.=-0.435, p=0.027) and the pallidum (est.=0.428, p=0.014) showed a significant predictive effect of z-score in relation to the precentral gyrus.

To explore the interaction between main effects, a two-factor interaction model (z-score ∼ diagnostic group:tracer + region + age + sex) was performed. The model was significant (F(45, 2783)=3.85, p<0.001), however, no interaction effect between diagnostic group and tracer was found (est.=0.130,p=0.136).

A three-factor interaction model (z-score ∼ diagnostic group:tracer:region + age + sex) was run to assess the interaction between diagnostic group and tracer for each region to predict z-score. This model was significant (F(165, 2663)=1.82, p<0.001), with the subcallosal area showing a significant interaction effect with diagnostic group and tracer (est.=-0.312, p=0.034).

Full output of the full factorial analyses can be found in supplementary table 1.

### Euclidean distance with hierarchical clustering

A Euclidean distance matrix was generated to visualise the pairwise dissimilarity between participants, based on z-scores of all brain regions. Figure 2 details the Euclidean distance matrices where hierarchical clustering of patients with PSP-RS (Figure 2a) and controls participants (Figure 2b) was employed. Supplementary figure 1 depicts Euclidean distance matrices where participants were clustered with K-means clustering.

**Fig. 2:**
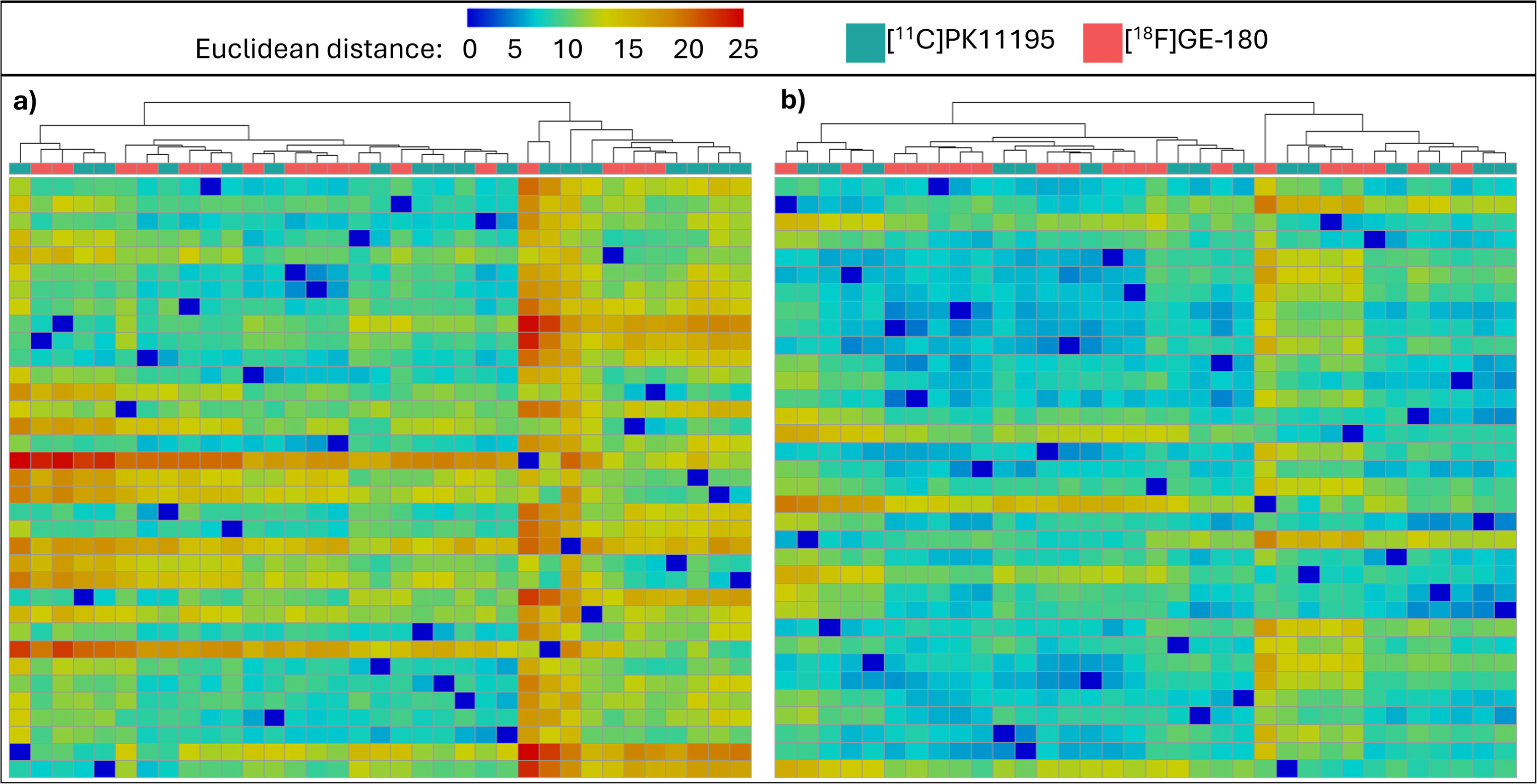
Clustering of Euclidean distance values based on z-scores of all brain regions. Each column of the heatmaps represent a participant, with the Euclidean distance value between each participant calculated based on all 41 brain regions. a) An agglomerative hierarchical clustering algorithm found that patients with PSP-RS spread evenly across the dendrogram despite tracer, suggesting homogeneity between tracers following harmonisation. b) An agglomerative hierarchical clustering algorithm also demonstrated an even spread of participants across the dendrogram in controls

### Representational similarity analysis with pattern similarity and permutation testing

Representational similarity analysis identified similar z-score patterns in patients with PSP-RS regardless of tracer (Figure 3). This was confirmed with pattern similarity between matrices (r=0.42) and permutation testing which demonstrated that it is highly unlikely this was a chance finding (Supplementary Figure 2). Although the relationship between z-score patterns in control participants was also significant (r=0.24) permutation testing highlighted that this finding could be due to chance (Supplementary Figure 1).

**Fig. 3:**
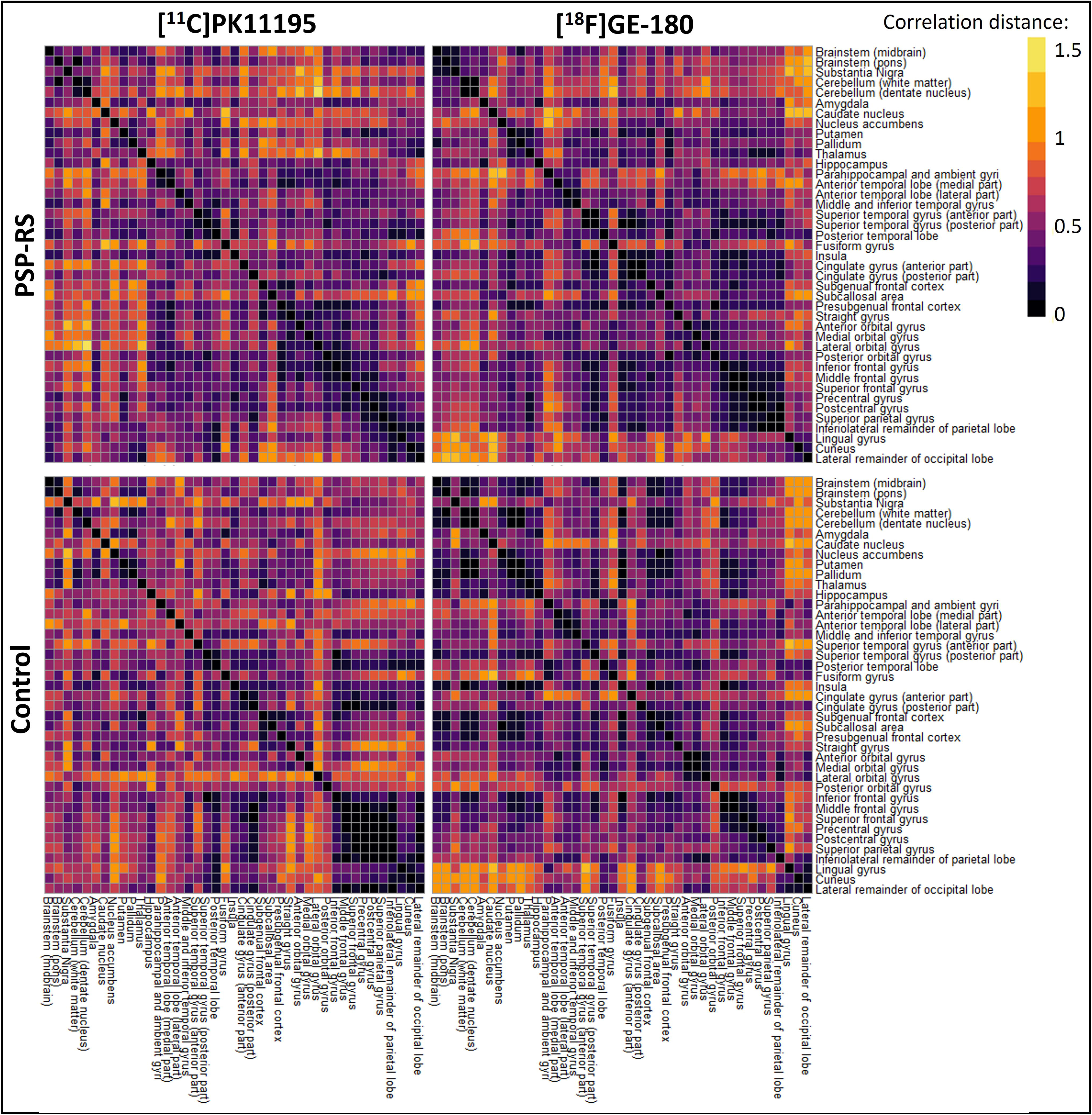
Representational similarity analysis. In patients with PSP-RS representational similarity matrices were visually similar, while matrices were less similar in controls. Pattern similarity between matrices and permutation testing are described in supplementary figure 2.

### Harmonisation across AD cohorts: [^11^C]PK11195, [^18^F]GE-180 and [^11^C]PBR28 results

Several dissimilarity analyses were run to determine whether any tracer-specific differences could be identified across the AD-specific cohorts.

### Pairwise comparisons

No differences in z-scores were observed between tracers for any brain region in control participants. In patients with AD, [^11^C]PK11195 had significantly greater binding than [^18^F]GE-180 in the pons (p=0.010) and pallidum (p=0.02), and significantly greater binding than [^11^C]PBR28 in the amygdala (p=0.002), medial anterior temporal lobe (p=0.04), and thalamus (p=0.03). [^18^F]GE-180 showed higher binding than [^11^C]PBR28 in the hippocampus (p=0.04) and amygdala (p=0.004), and higher binding than [^11^C]PK11195 in the subcallosal area (p=0.03). In the anterior cingulate, [^11^C]PBR28 demonstrated greater binding than [^18^F]GE-180 (p=0.03) (Figure 4).

**Fig. 4:**
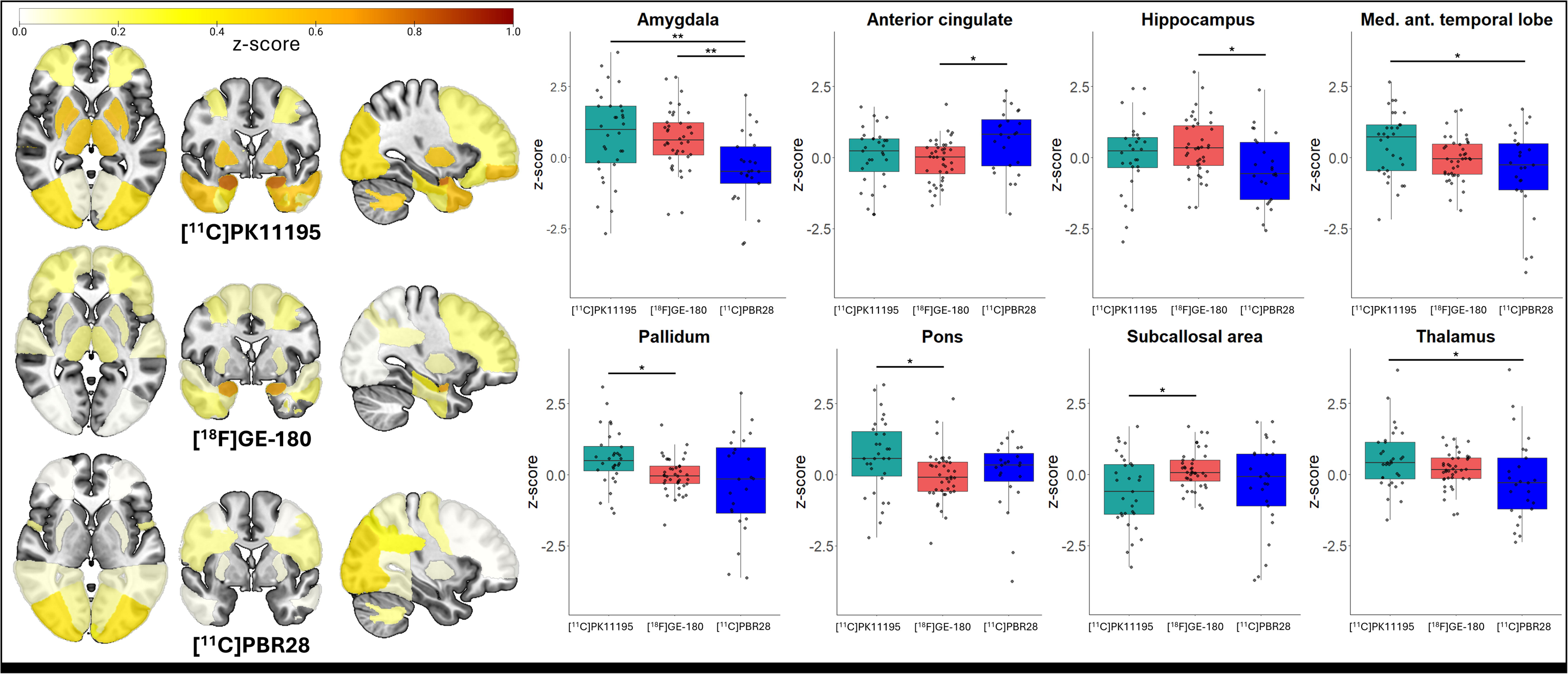
z-score brain plots of patients with AD. In patients with AD, elevated TSPO-PET z-scores were found across the brain compared to controls. 8 brain regions showed significant differences between tracers. *=p<0.05, **=p<0.001

### Full factorial analysis

The main effects model (z-score ∼ diagnostic group + tracer + region + age + sex) was significant F(45, 6350)=1.847, p<0.001), with a significant main effect seen when comparing [^11^C]PK11195 and [^11^C]PBR28 (est.=0.135, p=0.003) but not when comparing [^11^C]PK11195 and [^18^F]GE-180 or [^18^F]GE-180 and [^11^C]PBR28. While no main effect of age or sex was found, the amygdala (est.=0.301, p=0.037), caudate nucleus (est.=-0.381, p=0.008), and superior temporal gyrus anterior part (est.=-0.316, p=0.028) showed significant effects on the model.

To explore the interaction between main effects, a two-factor interaction model (z-score ∼ diagnostic group:tracer + region + age + sex) was performed and was significant (F(47, 6348)=1.91, p<0.001). Post-hoc analysis revealed that the difference between [^11^C]PK11195 and [^11^C]PBR28 in patients with AD (est.=0.222, p<0.001) was the main driver of the significance of this model.

A three-factor interaction model (z-score ∼ diagnostic group:tracer:region + age + sex) was run to assess the interaction between diagnostic group and tracer for each region to predict z-score. Overall, this model was not significant (F(247, 6148)=1.11, p=0.12), with the only significant interaction effect again being between [^11^C]PK11195 and [^11^C]PBR28 in the amygdala (est=1.50, p=0.048).

Full output of the full factorial analysis can be found in supplementary table 2.

### Euclidean distance with hierarchical clustering

A Euclidean distance matrix was generated to demonstrate the pairwise dissimilarity between AD and control participants, based on z-scores of all brain regions. Figure 5 details the Euclidean distance matrices where hierarchical clustering of patients with AD (Figure 5a) and control participants (Figure 5b) was employed. Supplementary figure 3 depicts Euclidean distance matrices with participants clustered using K-means clustering.

**Fig. 5:**
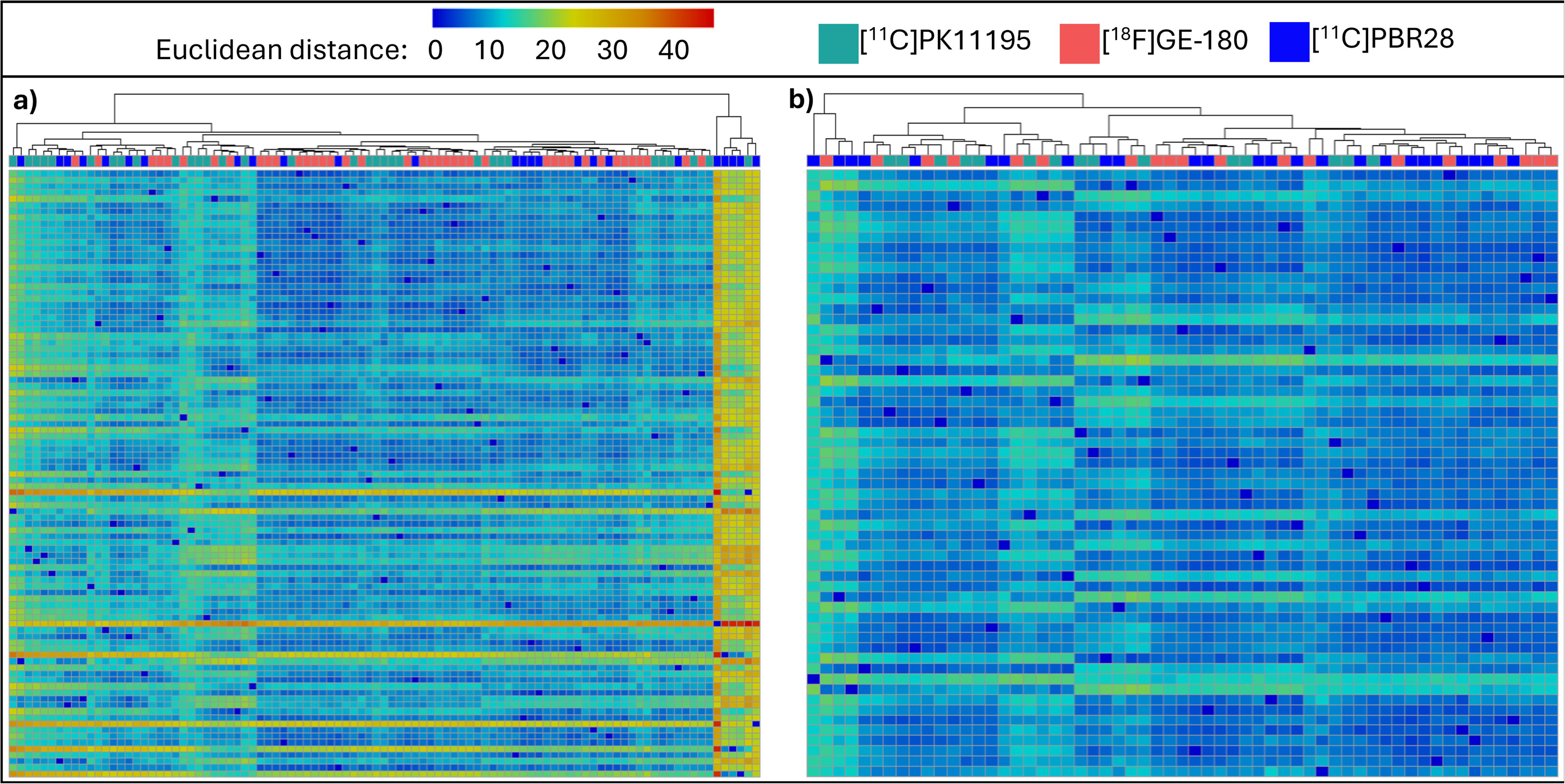
Clustering of Euclidean distance values based on z-scores of all brain regions. Each column of the heatmaps represent a participant, with the Euclidean distance value between each participant calculated based on all 41 brain regions. a) An agglomerative hierarchical clustering algorithm found a relatively even dispersion of patients with AD across the dendrogram, except for a small seperating cluster containing 5 [^11^C]PBR28 scanned patients and 1 [^11^C]PK11195 scanned patient. b) An agglomerative hierarchical clustering algorithm demonstrated an even spread of participants across the dendrogram in controls when all three tracers were included.

### Representational similarity analysis with pattern similarity and permutation testing

In the AD group, representational similarity analysis and pattern similarity of matrices identified similar z-score patterns between [^11^C]PK11195 and [^18^F]GE-180 (r=0.36), and between [^18^F]GE-180 and [^11^C]PBR28 (r=0.26) (Figure 6). Permutation testing of the weak correlation of patterns seen between [^11^C]PK11195 and [^11^C]PBR28 (r=0.15) (Figure 6) found that this was likely a chance finding (Supplementary Figure 4). Robust, however weak, correlations between tracers were seen in the control group; [^11^C]PK11195 and [^18^F]GE-180 (r=0.24), [^18^F]GE-180 and [^11^C]PBR28 (r=0.33), [^11^C]PK11195 and [^11^C]PBR28 (r=0.24) (Figure 6).

**Fig. 6:**
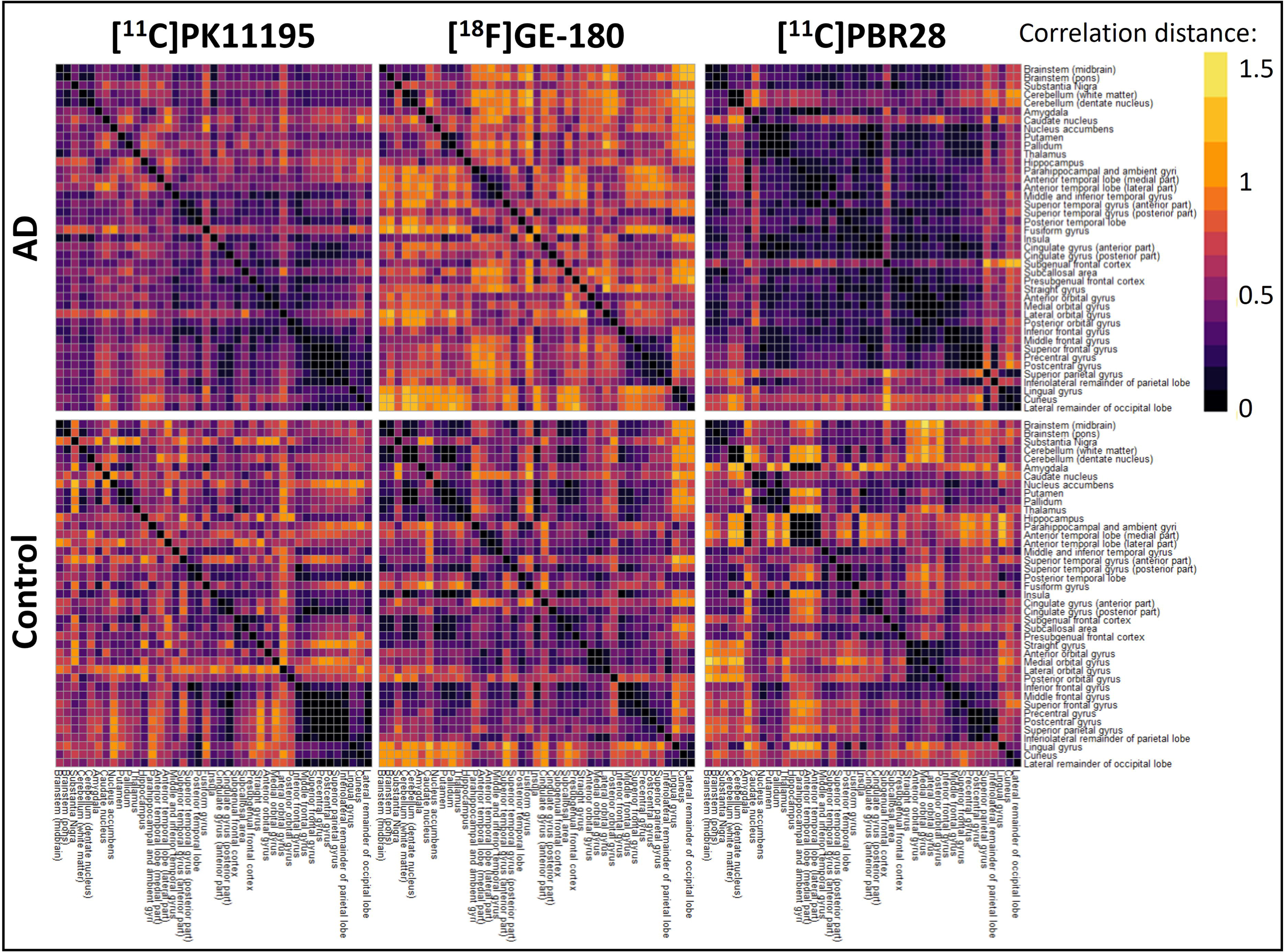
Representational similarity analysis. a) In patients with AD, the cohorts scanned with [^11^C]PK11195 and [^11^C]PBR28 demonstrated lower dissimilarity within groups compared to those scanned with [^18^F]GE-180. In control participants, no obvious pattern could be visualised from the dissimilarity matrices for any tracer. Supplementary Figure 4 depicts the pattern similarity analyses between matrices and permutation testing.

## Discussion

With the aim of harmonising and comparing TSPO-PET tracers across clinically matched cohorts of patients with primary and secondary tauopathies, we developed and applied a standardised pre-processing pipeline across three cohorts. We included patients with PSP-RS scanned with either [^11^C]PK11195 or [^18^F]GE-180, patients with AD scanned with either [^11^C]PK11195, [^18^F]GE-180, or [^11^C]PBR28, as well as control participants scanned with either of the three tracers. Overall, our multivariate approach and dissimilarity analyses converged to suggest that our new pipeline to harmonise TSPO-PET tracers and standardise the regional quantification of neuroinflammation is effective for [^11^C]PK11195 and [^18^F]GE-180 in PSP-RS. Due to its homogeneous and symmetrical neuropathological profile, PSP-RS is an excellent model disease to develop harmonisation and standardisation methodologies for TSPO-PET quantification across tracers. When applied to cohorts of patients with AD across three distinct tracers, the standardisation pipeline was less robust, with dissimilarity analyses highlighting that [^18^F]GE-180 and [^11^C]PK11195 were the most comparable, followed by [^18^F]GE-180 and [^11^C]PBR28, and lastly by [^11^C]PK11195 and [^11^C]PBR28.

With the emergence of novel, clinically meaningful, disease-modifying therapies for neurodegenerative diseases [41], efforts have been directed to harmonise methods that quantify the magnitude of neuropathological changes. For example, the Centiloid project [42] has revolutionised how amyloid-β pathology is evaluated based on a scale applicable to different amyloid-PET tracers, which allows for comparable amyloid measurements across tracers, cohorts and centres. This method has been particularly useful in trials of anti-amyloid therapies [43, 44]. Other efforts, such as the CenTauR [45] or Uni-Tau from HEAD studies [46] aim to standardise the quantification of tau pathology across tau-PET tracers and centres. While the interpretation of the TSPO-PET signal in humans remains a controversial topic, a recent study validated the signal using post-mortem tissue from patients with PSP who underwent TSPO-PET during life. Microglial levels of TSPO positively correlated with the ante-mortem TSPO-PET signal [4], reinforcing that this imaging tool provides a useful window to shed light on the levels of neuroinflammation that may be occurring within the brain. With increasing evidence on the key role of microglia-mediated inflammation in neurodegenerative diseases [47], and inflammation-targeting drugs under evaluation in several clinical trials [48], methods that enable harmonisation and combination of TSPO-PET tracers across centres are urgently needed to improve target-engagement evaluation, patient stratification and monitoring. Moving in this direction, recent work employed logistic regression to estimate the TSPO-PET signal across the brain using a dataset consisting of three tracers in healthy volunteers, patients with schizophrenia, AD, chronic pain, and in people given the TSPO blocking drug XBD173. Despite promising results, the high variability when extending the models to different tracers highlighted the need to optimise the model for each tracer [49]. Here, we focused on neurodegenerative diseases, specifically including patients with primary and secondary tauopathies. We have designed and implemented a standardised TSPO-PET processing pipeline to harmonise the pre-processing and post-processing analyses across TSPO-PET tracers in clinically matched cohorts, be able to compare and combine multi-centre data.

Using z-scores against centre- and tracer-specific controls, comparisons across brain regions demonstrated that tracers were comparable for 100% (41/41) of regions in control participants, 98% (40/41) of regions in patients with PSP-RS (Figure 1), and 80% (33/41) of regions in patients with AD (Figure 4). As z-scoring forced the mean of the control groups to zero, it was expected that no difference would be seen between tracers in this group. In patients with PSP-RS, tau deposition is usually predominant in subcortical regions with symmetrical patterns, before spreading throughout the cortex in later stages of the disease [50, 51]. Neuroinflammation has been shown to colocalise with and parallel the progression of tau pathology [15]. The high homogeneity of pathology seen in PSP-RS may be reflected in the similarities seen between tracers here, and highlight that the two tracers examined here can be compared in this disease group following harmonisation. In AD, although tau pathology usually follows Braak stages [52], the widespread presence of amyloid makes the neuroinflammatory profile much more heterogeneous in these patients compared to in PSP-RS [13, 26]. The heterogeneous nature of neuroinflammation in AD may be the reason why fewer regions were comparable between the three tracers, as patients with AD have more variable brain inflammation profiles. Furthermore, the AD cohort consisted of patients with either AD or MCI-AD, which may have resulted in the variability observed due to the greater range of disease severity in comparison to the PSP-RS group, which all had established disease and diagnoses.

We ran full factorial analyses to evaluate the main, two-, and three-factor interaction effects of tracer, diagnosis, brain region, age, and sex to predict TSPO-PET z-scores. This analysis demonstrated the presence of a main effect of tracers on the model in the PSP-RS cohort suggesting a difference between [^11^C]PK11195 and [^18^F]GE-180 in this group, however this effect only just reached significance (p=0.041) and may have been explained by other variables, such as diagnostic group, age, and sex. As expected, the diagnostic group variable, defined as PSP-RS vs control cohorts, showed a main effect on z-score prediction, as patients with PSP-RS generally have greater levels of neuroinflammation. Also reassuringly due to its impact in PSP-RS, the pallidum showed a significant predictive effect of z-score in the model. Significant interaction effects were identified when age and/or sex, and PSP-RS-effected regions, such as the subcallosal area, were included in interaction terms with the tracer variable. In the AD group, post-hoc analysis of the main effects model suggested that a significant effect was found when changing between [^11^C]PK11195 and [^11^C]PBR28, further highlighting that either tracer or patient-specific differences may be present in this group. Several interaction effects involving the tracer were also found. Sex and age consistently demonstrated significant main and interaction effects in these models, both in PSP-RS and AD. Sex differences in TSPO-PET binding have been identified in AD, with females showing greater binding than males [53]. App^NL-G-F^ Amyloidosis mouse models of AD replicate this finding but this is not seen in P301S mice exhibiting tau pathology [54]. It is now also largely proven that inflammation increases with age, with inflammaging being a risk factor for developing neurodegenerative disease [55]. However, further studies are needed to fully understand age- and sex-specific differences related to the TSPO signal and microglia-mediated cascades in tauopathies.

To further test our standardisation pipeline, Euclidean distances were calculated for each participant, based on all forty-one brain regions and clustered using two separate methods to visualise any tracer-specific clustering. Firstly, for patients with PSP-RS and controls, the hierarchical clustering algorithm demonstrated that individual inflammation patterns (i.e. single participants) were evenly distributed across the dendrogram, regardless of whether their TSPO levels were quantified with [^11^C]PK11195 or [^18^F]GE-180 (Figure 2). Next, K-means clustering with two clusters was used, and each data-driven cluster included participants from both tracer cohorts in both PSP-RS and control groups (Supplementary Figure 1), suggesting that the standardisation pipeline was effective. In patients with AD and controls scanned with [^11^C]PK11195, [^18^F]GE-180, or [^11^C]PBR28, again the control participants were evenly spread across the dendrogram when hierarchical clustering was used, and all three of the K-means clusters contained a mixture of all three tracers (Supplementary Figure 3). In the AD cohort, however, one small cluster containing five patients scanned with [^11^C]PBR28 and one with [^11^C]PK11195 clustered separately from all other participants, who were instead evenly spread across the rest of the matrix (Figure 5). These 6 patients had consistently low TSPO-PET z-scores compared to other participants throughout the brain; four of them were male and relatively unimpaired cognitively, with MMSE scores ≥ 24. As studies have demonstrated discrepancies in TSPO-PET binding as a result of factors such as sex (as well as age and body mass index) in both mice and humans [54, 56, 57], further work should aim to explore and understand individual differences in TSPO-PET binding in both healthy and diseased brains.

Lastly, a representational similarity analysis was run to visualise any regional z-score patterns between tracers. This was followed by a pattern similarity analysis of the representational similarity matrices to quantify the correlation between the tracers. As with previous analyses in the PSP-RS cohorts, [^11^C]PK11195 and [^18^F]GE-180 showed comparable z-score patterns across the brain, a result that permutation testing confirmed was robust (Supplementary Figure 2), which coincides with tau deposition [15], and further supports the comparability of these two tracers following implementation of the standardisation pipeline. In AD, a similarly strong and robust z-score pattern was seen for both [^11^C]PK11195 and [^18^F]GE-180, while [^18^F]GE-180 and [^11^C]PBR28 patterns were also statistically similar (Figure 6). Corroborating the results of the full factorial analysis, [^11^C]PK11195 and [^11^C]PBR28 regional z-scores patterns were not statistically similar in this AD cohort following standardisation, emphasising either tracer or cohort differences between these two groups. For control participants, weak but robust correlations in z-score patterns were seen between the three tracers, possibly highlighting a generalised pattern of background neuroinflammation ongoing in the brain to alleviate everyday insults, as well as confirming the absence of any specific neuropathology in these participants.

The differences observed between tracers here could be due to a number of factors: first, the differences may be tracer-specific, and our standardisation pipeline may need to be optimised for each tracer, as suggest by Maccioni and colleagues for their models [49]. Second, differences may also occur as a result of hardware discrepancies. As well as using different tracers, different PET scanners were used in Cambridge, Munich, and Montreal, which is an unavoidable difference that must be considered and appreciated in multi-centre imaging studies. Moving forward, a collaborative effort must address how best to account for differences in scanner outputs. Third, although clinically matched, the AD cohorts consisted of both patients with AD and patients with MCI-AD which may have increased the variation in neuroinflammation in these groups and affected their comparability. Lastly, neuroinflammation is a dynamic process, both in healthy and diseased brain states [6, 58, 59], therefore it is more difficult than for other pathological hallmark to fully match cohorts of people without scanning the same individuals with different tracers within a short timeframe. Indeed, head-to-head studies of TSPO-PET radiotracers have been completed, however these are limited to healthy volunteers only [30, 31]. It is therefore also likely that, especially in a heterogeneous disease like AD, individual patient differences could affect the performance of the standardisation pipeline. In addition to the increased variability possibly added by including patients with MCI-AD in the AD cohort, differences in PET scanners used at each centre may have introduced variation in the PET outputs and may have an inflated variability seen between tracers.

Together, our work suggests that harmonisation pipelines for TSPO-PET tracers can be applied for PSP-RS, likely due to the homogeneous and symmetrical neuropathological profile of PSP-RS rendering it an excellent model disease to develop harmonisation and standardisation methodologies for TSPO-PET quantification across tracers. We found, however, that when applied to patients with AD across three distinct tracers, our standardisation pipeline was less robust, with dissimilarity analyses highlighting that [^18^F]GE-180 and [^11^C]PK11195 were the most comparable, followed by [^18^F]GE-180 and [^11^C]PBR28, and lastly by [^11^C]PK11195 and [^11^C]PBR28. Ongoing work aims to further optimise and scrutinise standardisation pipelines, whilst expanding the approach to larger cohorts, a greater number of tracers, and other neurodegenerative diseases. It will be important to validate and expand this work to develop thresholds of “neuroinflammation severity” using multiple TSPO-PET tracers related to clinical and cognitive outcomes in order to allow for the creation of multi-centre databases, larger than those which could be created at a single site. The final goal is to have powerful TSPO-PET studies, where large cohorts can be analysed and compared quantitatively to inform clinical trials targeting inflammation and provide an objective and reliable outcome and/or monitoring measure.

## Ethics approval

This study was performed in line with the principles of the Declaration of Helsinki. The following ethics approvals were granted: The research protocol for Cambridge data were approved by the National Research Ethics Service’s East of England Cambridge Central Committee, and the UK Administration of Radioactive Substances Advisory Committee. The study protocol for LMU Munich data was approved by the Ethics Committee of LMU Munich and the Bundesamt für Strahlenschutz (project numbers 17-755 and 17-569).

The protocol for the TRIAD study was approved by the Montreal Neurological Institute PET working committee and the Douglas Mental Health University Institute Research Ethics Board.

## Acknowledgments

We thank our participant volunteers for their participation in this study, the clinical staff and the research nurses for their contribution, and the East Anglia Dementias and Neurodegenerative Diseases Research Network (DeNDRoN) for help with subject recruitment. We thank all our patients, their caregivers, cyclotron, radiochemistry, and the positron emission tomography imaging staff at LMU Munich. We thank all of our participants and their care partners who contributed to this work at the Montreal Neurological Institute, McGill University.

## Declarations

### Acknowledgments

We thank our participant volunteers for their participation in this study, the clinical staff and the research nurses for their contribution, and the East Anglia Dementias and Neurodegenerative Diseases Research Network (DeNDRoN) for help with subject recruitment. We thank the UK Dementia Research Institute (UKDRI) at Cambridge. We thank all our patients, their caregivers, cyclotron, radiochemistry, and the positron emission tomography imaging staff at LMU Munich. GE Healthcare made GE-180 cassettes available through an early access model. We thank all of our participants and their care partners who contributed to this work at the Montreal Neurological Institute, McGill University.

For the purpose of open access, the authors have applied a Creative Commons Attribution (CC BY) license to any Author Accepted Manuscript version arising from this submission.

### Funding

Cambridge University: This study was co-funded by the Dementias Platform UK and Medical Research Council (MC_UU_00030/14; MR/T033371/1); Race Against Dementia Alzheimer’s Research UK (ARUK-RADF2021A-010); Alzheimer’s Research UK PhD Scholarship (ARUK-PhD2023-018); the Wellcome trust (220258); the Cambridge University Centre for Parkinson-Plus (RG95450); the National Institute for Health Research (NIHR) Cambridge Biomedical Research Centre (NIHR203312: the views expressed are those of the authors and not necessarily those of the NIHR or the Department of Health and Social Care). This work is also supported by the UK Dementia Research Institute through UK DRI ltd, principally funded by the Medical Research Council.

LMU Munich: This study was supported by the German Center for Neurodegenerative Disorders (Deutsches Zentrum für Neurodegenerative Erkrankungen), Hirnliga (Manfred-Strohscheer Stiftung), and the German Research Foundation (Deutsche Forschungsgemeinschaft) under Germany’s Excellence Strategy within the framework of the Munich Cluster for Systems Neurology (EXC 2145 SyNergy, ID 390857198).

McGill University: The TRIAD cohort study data is supported by the Weston Brain Institute, Canadian Institutes of Health Research (CIHR) [MOP-11-51-31; RFN 152985, 159815, 162303], Canadian Consortium of Neurodegeneration and Aging (CCNA; MOP-11-51-31-team 1), Brain Canada Foundation (CFI Project 34874; 33397), the Fonds de Recherche du Québec – Santé (FRQS; 2020-VICO-279314; 2024 VICO-356138; https://doi.org/10.69777/324345; https://doi.org/10.69777/356138 and https://doi.org/10.69777/312994), and the Colin J Adair Charitable Foundation.

### Competing Interests

The authors have no conflicts of interest to report related to this work. Unrelated to this work, JTO has received honoraria unrelated to this work as DSMB chair or member for TauRx, Axon, Eisai and Novo Nordisk, and has acted as a consultant for Biogen and Roche, and has received research support from Alliance Medical and Merck. JBR is a non-remunerated trustee of the Guarantors of Brain, Darwin College and the PSP Association (UK). He provides consultancy unrelated to the current work to Asceneuron, Astronautx, Astex, Alector, Booster Therapeutics, Curasen, CumulusNeuro, ClinicalInk, Draig Therapeutics, Eisai, Ferrer, Wave, SVHealth, and has research grants from AZ-Medimmune, Janssen, and Lilly as industry partners in the Dementias Platform UK. MM has acted as a consultant for Astex Pharmaceuticals. RP has received honoraria for advisory boards and speaker engagements from Roche, EISAI, Eli Lilly, Biogen, Janssen-Cilag, AstraZeneca, Schwabe, Grifols, Novo Nordisk, and Tabuk, and is a shareholder of Medotrax GmbH and Vistim Ltd. MB received speaker honoraria from GE healthcare, Roche, Iba, and Life Molecular Imaging and is an advisor of GE healthcare, MIAC, and Life Molecular Imaging. NF received speaker or consulting honoraria from GE healthcare, EISAI, Life molecular imaging and MSD.

### Authors Contributions

MM, MB, and PRN were responsible for study conception and design. Patient recruitment, material preparation, and data collection were performed by NF, NR, JSG, AS, SNRC, CP, PSJ, TF, YH, FIA, RH, BSR, GM, JL, GH, JBR, RP, and JTO. Data analyses were performed by HC, NF, NR, JSG, TF, and YH. The first draft of the manuscript was written by HC and all authors commented on the previous versions of the manuscript. All authors read and approved the final manuscript.

### Data Availability

Anonymized processed data can be shared upon request to the corresponding or senior authors. Raw data may also be requested but are likely to be subject to a data transfer agreement with restrictions required to comply with participant consent and data protection regulations.

### Consent to participate

Participants with mental capacity gave their written informed consent to take part in the study. For those who lacked capacity, their participation followed the consultee process in accordance with the law.

### Consent to publish

By providing written informed consent to take part in the study, participants consented to anonymised results being published.

## Notes

### Author Declarations

The following ethics approvals were granted: The research protocol for Cambridge data were approved by the National Research Ethics Services East of England Cambridge Central Committee, and the UK Administration of Radioactive Substances Advisory Committee. The study protocol for LMU Munich data was approved by the Ethics Committee of LMU Munich and the Bundesamt fur Strahlenschutz (project numbers 17-755 and 17-569). The protocol for the TRIAD study was approved by the Montreal Neurological Institute PET working committee and the Douglas Mental Health University Institute Research Ethics Board.

