## Supplementary figures and images for "Comparing and combining TSPO-PET tracers in tauopathies"

### supplementary figure 1

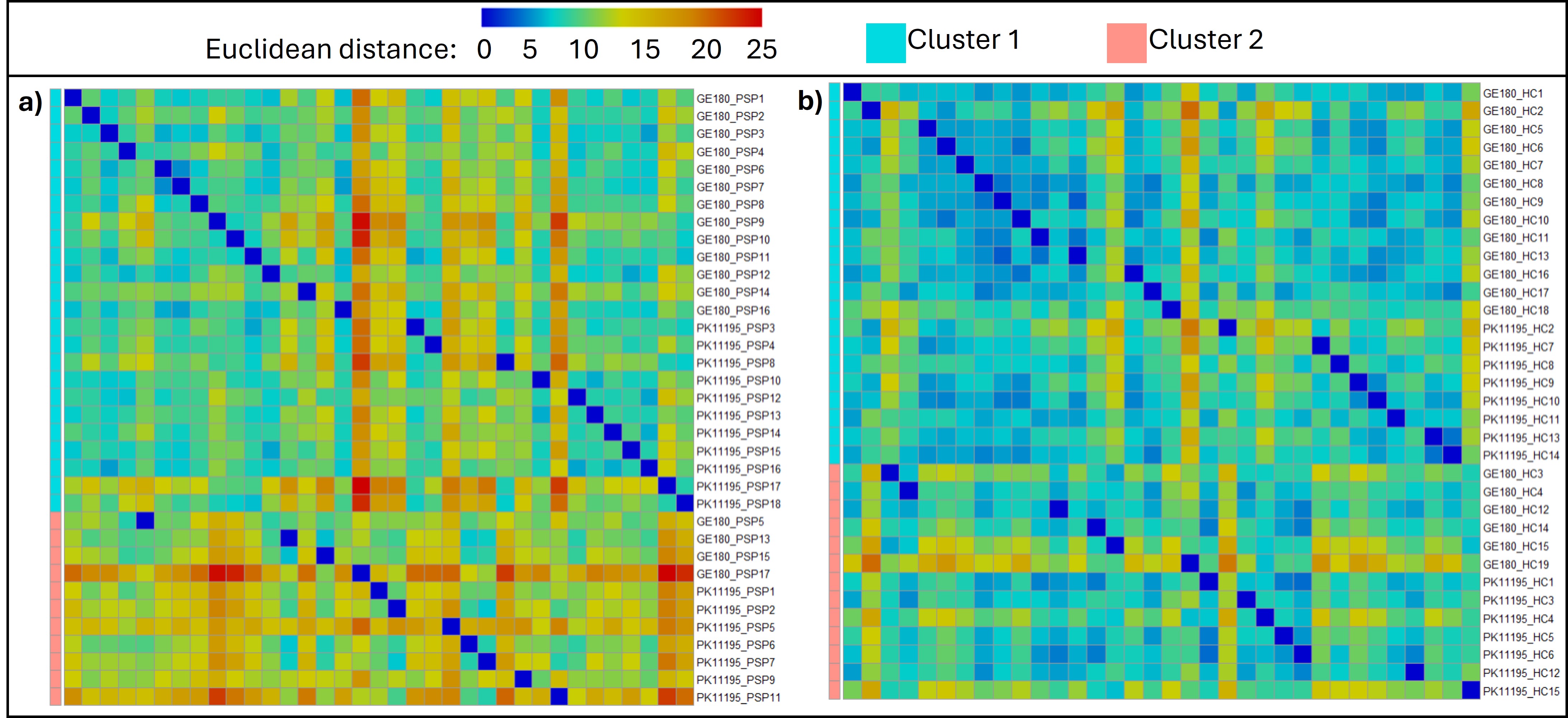

### supplementary figure 2

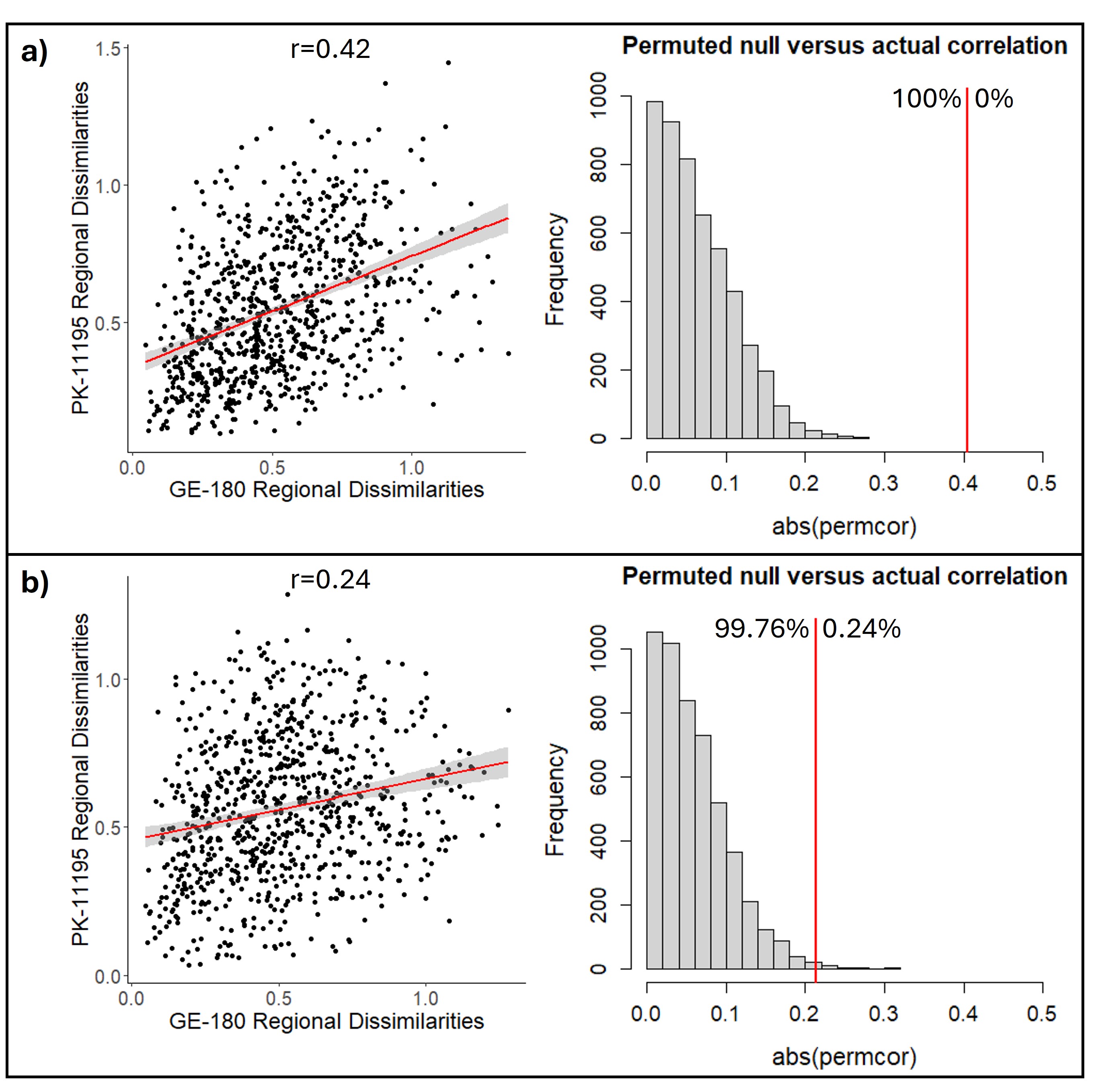

### supplementary figure 4

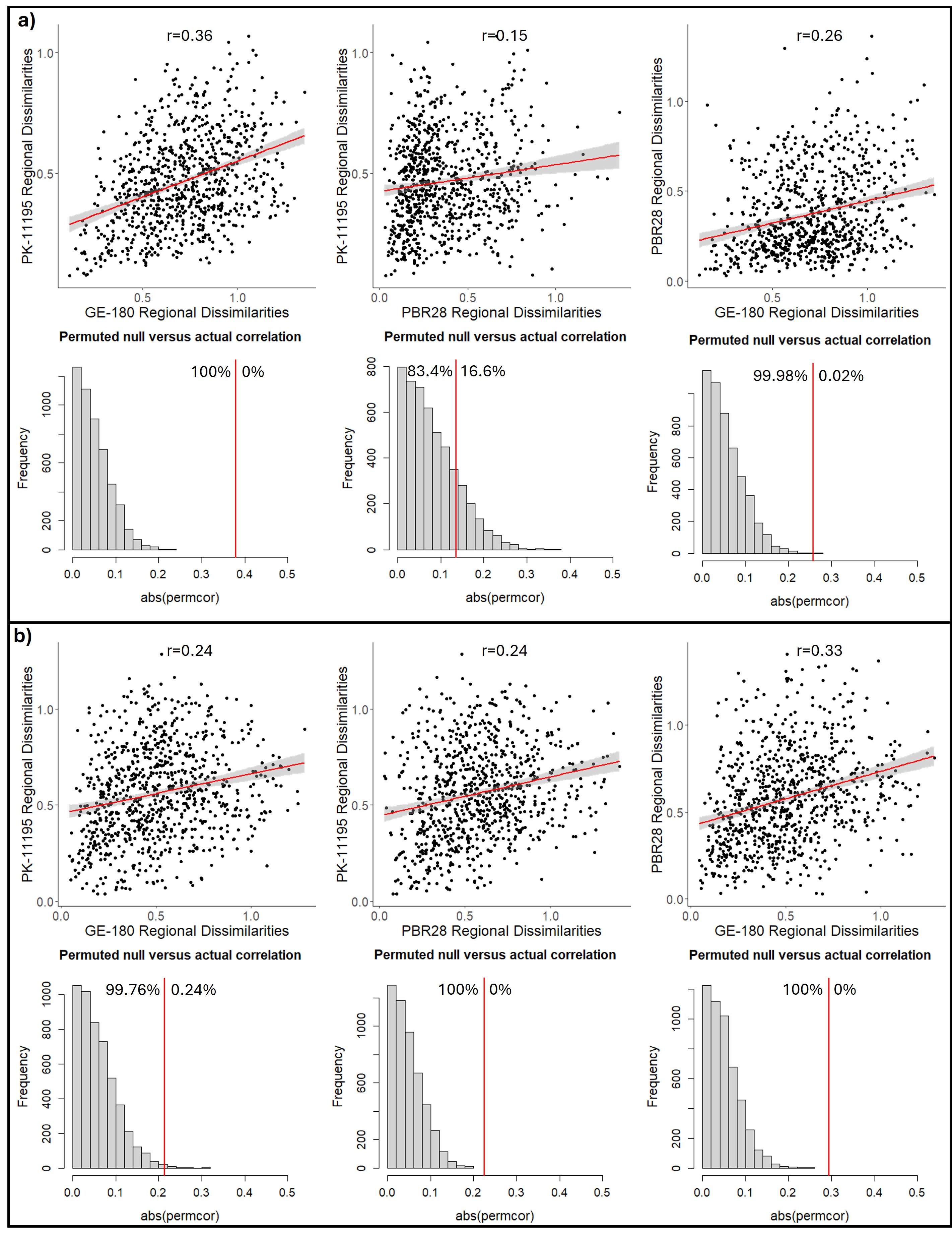

### supplementary table 3

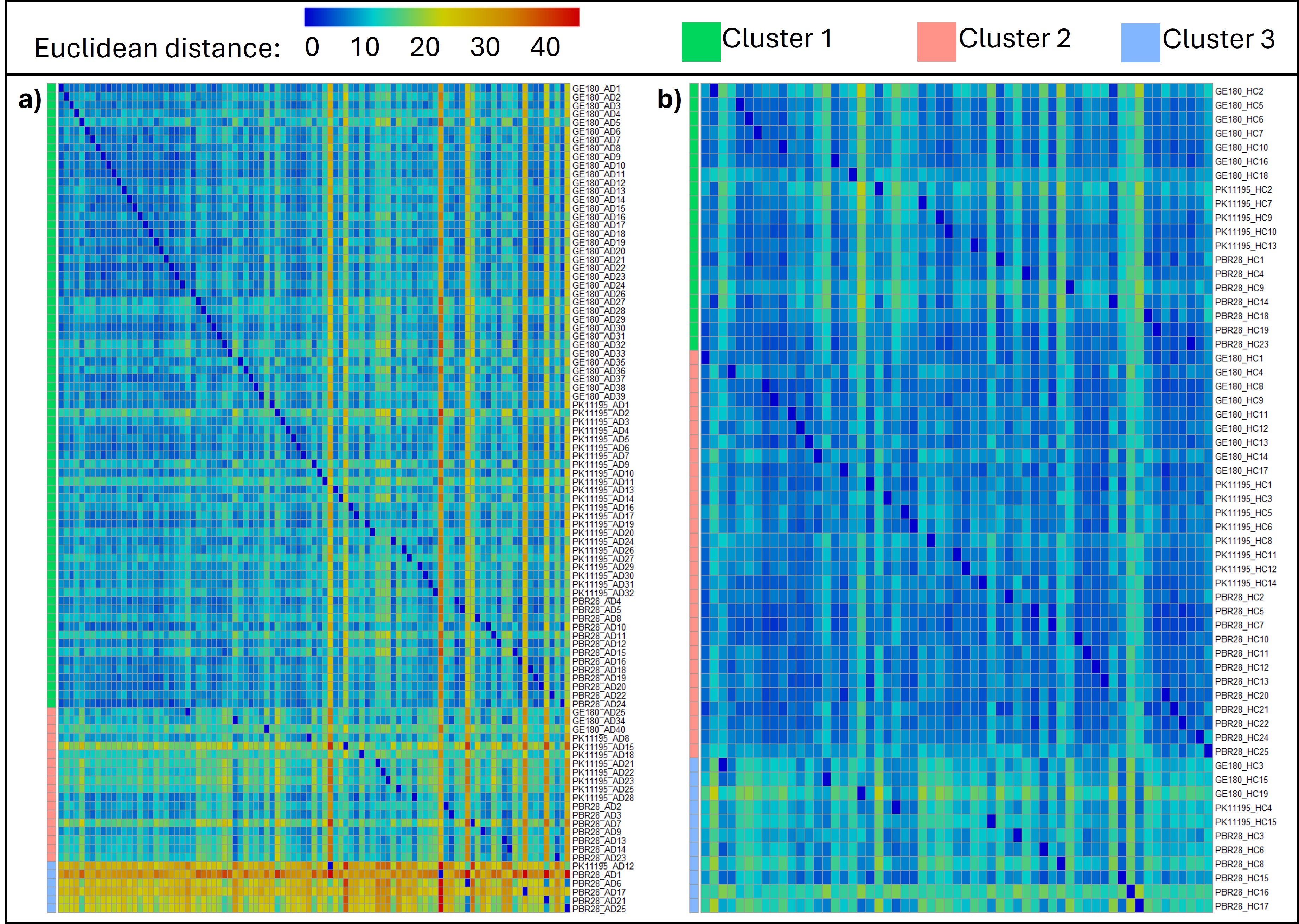
