## supplementary figure 1 for "Comparing and combining TSPO-PET tracers in tauopathies"

c) When forced into 2 clusters through K-means clustering, both clusters contained a mix of patients with PSP-RS scanned with either [^11^C]PK11195 or [^18^F]GE-180. d) When forced into 2 clusters through K-means clustering, both clusters contained a mix of controls scanned with either [^11^C]PK11195 or [^18^F]GE-180.

Supplementary figure 2:

Supplementary figure 3:

c) When forced into 3 clusters through K-means clustering, the same 5 [^11^C]PBR28 and 1 [^11^C]PK11195 scanned patients with AD formed a separate cluster, while the other two clusters contained a mix of patients scanned with each tracer. d) When forced into 3 clusters through K-means clustering, all clusters contained a mix of controls scanned with either [^11^C]PK11195, [^18^F]GE-180, or [^11^C]PBR28.

Supplementary figure 4:
