## supplementary table 1 for "Comparing and combining TSPO-PET tracers in tauopathies"

**Supplementary table 1: Outputs of main, two- and three- factor interaction models in the PSP-RS cohort.**

| **Main effects model (z-score ~ diagnostic group + tracer + region + age + sex** | | | | |
| --- | --- | --- | --- | --- |
| **Contrast** | **Estimate** | **Standard error** | **t ratio** | **p** |
| Diagnostic group | 0.136 | 0.0443 | 3.06 | 0.002 |
| Tracer | 0.0893 | 0.0438 | 2.04 | 0.041 |
| Precentral gyrus | ref | ref | *ref* | *ref* |
| Amygdala | 0.0541 | 0.197 | 0.275 | 0.783 |
| Anterior temporal lobe lateral part | -0.128 | 0.197 | -0.649 | 0.517 |
| Anterior temporal lobe medial part | -0.165 | 0.197 | -0.838 | 0.402 |
| Midbrain | 0.232 | 0.197 | 1.18 | 0.238 |
| Pons | 0.0265 | 0.197 | 0.135 | 0.893 |
| Caudate nucleus | -0.270 | 0.197 | -1.37 | 0.171 |
| Cerebellum dentate | 0.197 | 0.197 | 1.001 | 0.317 |
| Cerebellum white matter | 0.0781 | 0.197 | 0.397 | 0.692 |
| Inferior frontal gyrus | 0.0708 | 0.197 | 0.359 | 0.719 |
| Middle frontal gyrus | 0.0993 | 0.197 | 0.504 | 0.614 |
| Anterior orbital gyrus | 0.337 | 0.197 | 1.71 | 0.088 |
| Lateral orbital gyrus | 0.286 | 0.197 | 1.45 | 0.146 |
| Medial orbital gyrus | 0.173 | 0.197 | 0.879 | 0.379 |
| Posterior orbital gyrus | 0.0765 | 0.197 | 0.388 | 0.698 |
| Straight gyrus | 0.0260 | 0.197 | 0.132 | 0.895 |
| Superior frontal gyrus | -0.0518 | 0.197 | -0.263 | 0.792 |
| Cingulate gyrus anterior part | -0.0549 | 0.197 | -0.279 | 0.781 |
| Cingulate gyrus posterior part | -0.0562 | 0.197 | -0.286 | 0.775 |
| Fusiform gyrus | 0.239 | 0.197 | 1.21 | 0.226 |
| Parahippocampal and ambient gyri | 0.0304 | 0.197 | 0.154 | 0.877 |
| Superior temporal gyrus anterior part | -0.435 | 0.197 | -2.21 | 0.027 |
| Superior temporal gyrus posterior part | -0.0825 | 0.197 | -0.419 | 0.675 |
| Middle and inferior temporal gyrus | 0.0657 | 0.197 | 0.334 | 0.739 |
| Hippocampus | -0.0969 | 0.197 | -0.492 | 0.623 |
| Insula | 0.0482 | 0.197 | 0.245 | 0.807 |
| Nucleus accumbens | -0.0892 | 0.197 | -0.453 | 0.651 |
| Cuneus | -0.130 | 0.197 | -0.659 | 0.510 |
| Lingual gyrus | -0.181 | 0.197 | -0.918 | 0.359 |
| Lateral remainder of the occipital lobe | -0.0787 | 0.197 | -0.4 | 0.689 |
| Pallidum | 0.483 | 0.197 | 2.45 | 0.014 |
| Postcentral gyrus | -0.148 | 0.197 | -0.751 | 0.453 |
| Inferiolateral remainder of parietal lobe | -0.0591 | 0.197 | -0.3 | 0.764 |
| Superior parietal gyrus | -0.116 | 0.197 | -0.591 | 0.555 |
| Posterior temporal lobe | -0.147 | 0.197 | -0.748 | 0.455 |
| Presubgenual frontal cortex | -0.00583 | 0.197 | -0.03 | 0.976 |
| Putamen | 0.204 | 0.197 | 1.04 | 0.299 |
| Substantia nigra | 0.0320 | 0.197 | 0.162 | 0.871 |
| Subcallosal area | -0.214 | 0.197 | -1.09 | 0.278 |
| Subgenual frontal cortex | -0.0559 | 0.197 | -0.284 | 0.776 |
| Thalamus | 0.133 | 0.197 | 0.676 | 0.499 |
| Age | 0.0327 | 0.00335 | 9.76 | <0.001 |
| Sex | -0.245 | 0.0454 | -5.39 | <0.001 |
| **Two factor interaction model (z-score ~ diagnostic group:tracer + region + age + sex)** | | | | |
| **Contrast** | **Estimate** | **Standard error** | **t ratio** | **p** |
| Diagnostic group:Tracer | 0.130 | 0.0874 | 1.49 | 0.136 |
| Diagnostic group | 0.0738 | 0.0606 | 1.22 | 0.224 |
| Tracer | 0.0228 | 0.0625 | 0.365 | 0.715 |
| Precentral gyrus | ref | ref | ref | ref |
| Amygdala | 0.0541 | 0.197 | 0.275 | 0.783 |
| Anterior temporal lobe lateral part | -0.128 | 0.197 | -0.649 | 0.516 |
| Anterior temporal lobe medial part | -0.165 | 0.197 | -0.838 | 0.402 |
| Midbrain | 0.232 | 0.197 | 1.18 | 0.238 |
| Pons | 0.0265 | 0.197 | 0.135 | 0.893 |
| Caudate nucleus | -0.270 | 0.197 | -1.37 | 0.171 |
| Cerebellum dentate | 0.197 | 0.197 | 1.00 | 0.317 |
| Cerebellum white matter | 0.0781 | 0.197 | 0.397 | 0.692 |
| Inferior frontal gyrus | 0.0708 | 0.197 | 0.360 | 0.719 |
| Middle frontal gyrus | 0.0993 | 0.197 | 0.504 | 0.614 |
| Anterior orbital gyrus | 0.337 | 0.197 | 1.71 | 0.0874 |
| Lateral orbital gyrus | 0.286 | 0.197 | 1.45 | 0.146 |
| Medial orbital gyrus | 0.173 | 0.197 | 0.879 | 0.380 |
| Posterior orbital gyrus | 0.0765 | 0.197 | 0.388 | 0.698 |
| Straight gyrus | 0.0260 | 0.197 | 0.132 | 0.895 |
| Superior frontal gyrus | -0.0518 | 0.197 | -0.263 | 0.792 |
| Cingulate gyrus anterior part | -0.0549 | 0.197 | -0.279 | 0.781 |
| Cingulate gyrus posterior part | -0.0562 | 0.197 | -0.286 | 0.775 |
| Fusiform gyrus | 0.239 | 0.197 | 1.21 | 0.225 |
| Parahippocampal and ambient gyri | 0.0304 | 0.197 | 0.154 | 0.877 |
| Superior temporal gyrus anterior part | -0.435 | 0.197 | -2.21 | 0.0271 |
| Superior temporal gyrus posterior part | -0.0825 | 0.197 | -0.419 | 0.675 |
| Middle and inferior temporal gyrus | 0.0657 | 0.197 | 0.334 | 0.739 |
| Hippocampus | -0.0969 | 0.197 | -0.492 | 0.622 |
| Insula | 0.0482 | 0.197 | 0.245 | 0.807 |
| Nucleus accumbens | -0.0892 | 0.197 | -0.453 | 0.650 |
| Cuneus | -0.130 | 0.197 | -0.659 | 0.510 |
| Lingual gyrus | -0.181 | 0.197 | -0.918 | 0.359 |
| Lateral remainder of the occipital lobe | -0.0787 | 0.197 | -0.400 | 0.689 |
| Pallidum | 0.483 | 0.197 | 2.45 | 0.0143 |
| Postcentral gyrus | -0.148 | 0.197 | -0.751 | 0.453 |
| Inferiolateral remainder of parietal lobe | -0.0591 | 0.197 | -0.300 | 0.764 |
| Superior parietal gyrus | -0.116 | 0.197 | -0.591 | 0.555 |
| Posterior temporal lobe | -0.147 | 0.197 | -0.748 | 0.454 |
| Presubgenual frontal cortex | -0.00583 | 0.197 | -0.0300 | 0.976 |
| Putamen | 0.204 | 0.197 | 1.04 | 0.300 |
| Substantia nigra | 0.0320 | 0.197 | 0.162 | 0.871 |
| Subcallosal area | -0.214 | 0.197 | -1.09 | 0.278 |
| Subgenual frontal cortex | -0.0559 | 0.197 | -0.284 | 0.776 |
| Thalamus | 0.133 | 0.197 | 0.676 | 0.499 |
| Age | 0.0327 | 0.00335 | 9.77 | <0.001 |
| Sex | -0.248 | 0.0454 | -5.45 | <0.001 |
| **Three factor interaction model (z-score ~ diagnostic group:tracer:region + age + sex)** | | | | |
| **Contrast** | **Estimate** | **Standard error** | **t ratio** | **p** |
| Diagnostic group:Tracer:Precentral gyrus | *ref* | *ref* | *ref* | *ref* |
| Diagnostic group:Tracer:Amygdala | -0.17 | 0.79 | -0.218 | 0.8273 |
| Diagnostic group:Tracer:Anterior temporal lobe lateral part | 0.28 | 0.79 | 0.355 | 0.7225 |
| Diagnostic group:Tracer:Anterior temporal lobe medial part | -0.37 | 0.79 | -0.469 | 0.6394 |
| Diagnostic group:Tracer:Midbrain | -0.77 | 0.79 | -0.981 | 0.3268 |
| Diagnostic group:Tracer:Pons | -0.43 | 0.79 | -0.547 | 0.5847 |
| Diagnostic group:Tracer:Caudate nucleus | -0.40 | 0.79 | -0.507 | 0.6123 |
| Diagnostic group:Tracer:Cerebellum dentate | 0.10 | 0.79 | 0.131 | 0.8961 |
| Diagnostic group:Tracer:Cerebellum white matter | -0.25 | 0.79 | -0.323 | 0.747 |
| Diagnostic group:Tracer:Inferior frontal gyrus | -0.12 | 0.79 | -0.156 | 0.8761 |
| Diagnostic group:Tracer:Middle frontal gyrus | -0.09 | 0.79 | -0.112 | 0.9106 |
| Diagnostic group:Tracer:Anterior orbital gyrus | 0.80 | 0.79 | 1.014 | 0.3107 |
| Diagnostic group:Tracer:Lateral orbital gyrus | 0.34 | 0.79 | 0.426 | 0.6703 |
| Diagnostic group:Tracer:Medial orbital gyrus | -0.03 | 0.79 | -0.04 | 0.9679 |
| Diagnostic group:Tracer:Posterior orbital gyrus | 0.09 | 0.79 | 0.116 | 0.9074 |
| Diagnostic group:Tracer:Straight gyrus | -0.77 | 0.79 | -0.974 | 0.3302 |
| Diagnostic group:Tracer:Superior frontal gyrus | -0.43 | 0.79 | -0.541 | 0.5888 |
| Diagnostic group:Tracer:Cingulate gyrus anterior part | -0.68 | 0.79 | -0.864 | 0.3879 |
| Diagnostic group:Tracer:Cingulate gyrus posterior part | -0.16 | 0.79 | -0.203 | 0.8388 |
| Diagnostic group:Tracer:Fusiform gyrus | 0.65 | 0.79 | 0.821 | 0.4119 |
| Diagnostic group:Tracer:Parahippocampal and ambient gyri | 0.31 | 0.79 | 0.391 | 0.6959 |
| Diagnostic group:Tracer:Superior temporal gyrus anterior part | -0.95 | 0.79 | -1.199 | 0.2305 |
| Diagnostic group:Tracer:Superior temporal gyrus posterior part | -0.69 | 0.79 | -0.871 | 0.3841 |
| Diagnostic group:Tracer:Middle and inferior temporal gyrus | 0.50 | 0.79 | 0.63 | 0.5286 |
| Diagnostic group:Tracer:Hippocampus | -0.53 | 0.79 | -0.674 | 0.5003 |
| Diagnostic group:Tracer:Insula | -0.48 | 0.79 | -0.605 | 0.5454 |
| Diagnostic group:Tracer:Nucleus accumbens | -0.51 | 0.79 | -0.648 | 0.517 |
| Diagnostic group:Tracer:Cuneus | 0.60 | 0.79 | 0.765 | 0.4445 |
| Diagnostic group:Tracer:Lingual gyrus | 0.36 | 0.79 | 0.459 | 0.6461 |
| Diagnostic group:Tracer:Lateral remainder of the occipital lobe | 0.58 | 0.79 | 0.73 | 0.4652 |
| Diagnostic group:Tracer:Pallidum | 0.26 | 0.79 | 0.327 | 0.7435 |
| Diagnostic group:Tracer:Postcentral gyrus | 0.14 | 0.79 | 0.176 | 0.8599 |
| Diagnostic group:Tracer:Inferiolateral remainder of parietal lobe | 0.20 | 0.79 | 0.252 | 0.8014 |
| Diagnostic group:Tracer:Superior parietal gyrus | 0.28 | 0.79 | 0.355 | 0.7224 |
| Diagnostic group:Tracer:Posterior temporal lobe | 0.18 | 0.79 | 0.224 | 0.8229 |
| Diagnostic group:Tracer:Presubgenual frontal cortex | -0.20 | 0.79 | -0.248 | 0.8043 |
| Diagnostic group:Tracer:Putamen | -0.03 | 0.79 | -0.034 | 0.9726 |
| Diagnostic group:Tracer:Substantia nigra | -0.06 | 0.79 | -0.08 | 0.9359 |
| Diagnostic group:Tracer:Subcallosal area | -1.68 | 0.79 | -2.122 | 0.0339 |
| Diagnostic group:Tracer:Subgenual frontal cortex | -0.32 | 0.79 | -0.401 | 0.6882 |
| Diagnostic group:Tracer:Thalamus | -0.60 | 0.79 | -0.762 | 0.4459 |
