## supplementary table 2 for "Comparing and combining TSPO-PET tracers in tauopathies"

**Supplementary table 2: Outputs of main, two- and three- factor interaction models and post-hoc analysis in the AD cohort.**

| **Main effects model (z-score ~ diagnostic group + tracer + region + age + sex** | | | | |
| --- | --- | --- | --- | --- |
| **Contrast** | **Estimate** | **Standard error** | **t ratio** | **p** |
| Diagnostic group | 0.135 | 0.0333 | 1.02 | 0.310 |
| Tracer ([^11^C]PK11195 – [^18^F]GE-180) | 0.0677 | 0.0390 | 1.72 | 0.199 |
| Tracer ([^11^C]PK11195 – [^11^FC]PBR28) | 0.301 | 0.0409 | 3.29 | 0.00290 |
| Tracer ([^18^F]GE-180 – [^11^FC]PBR28) | -0.0682 | 0.0387 | 1.75 | 0.^18^7 |
| Precentral gyrus | *ref* | *ref* | *ref* | *ref* |
| Amygdala | 0.0675 | 0.144 | 2.09 | 0.0366 |
| Anterior temporal lobe lateral part | 0.109 | 0.144 | -0.474 | 0.635 |
| Anterior temporal lobe medial part | 0.123 | 0.144 | 0.470 | 0.639 |
| Midbrain | -0.381 | 0.144 | 0.757 | 0.449 |
| Pons | 0.217 | 0.144 | 0.857 | 0.392 |
| Caudate nucleus | 0.158 | 0.144 | -2.65 | 0.00804 |
| Cerebellum dentate | 0.0166 | 0.144 | 1.51 | 0.131 |
| Cerebellum white matter | 0.132 | 0.144 | 1.10 | 0.272 |
| Inferior frontal gyrus | 0.141 | 0.144 | 0.^11^5 | 0.908 |
| Middle frontal gyrus | -0.00^11^7 | 0.144 | 0.921 | 0.357 |
| Anterior orbital gyrus | -0.0170 | 0.144 | 0.984 | 0.325 |
| Lateral orbital gyrus | -0.0316 | 0.144 | -0.00800 | 0.993 |
| Medial orbital gyrus | -0.0861 | 0.144 | -0.^118^ | 0.906 |
| Posterior orbital gyrus | -0.0133 | 0.144 | -0.220 | 0.826 |
| Straight gyrus | -0.129 | 0.144 | -0.599 | 0.549 |
| Superior frontal gyrus | 0.0978 | 0.144 | -0.0930 | 0.926 |
| Cingulate gyrus anterior part | 0.0882 | 0.144 | -0.897 | 0.370 |
| Cingulate gyrus posterior part | 0.120 | 0.144 | 0.680 | 0.496 |
| Fusiform gyrus | -0.316 | 0.144 | 0.614 | 0.539 |
| Parahippocampal and ambient gyri | -0.0623 | 0.144 | 0.832 | 0.405 |
| Superior temporal gyrus anterior part | 0.2^11^ | 0.144 | -2.20 | 0.0280 |
| Superior temporal gyrus posterior part | -0.0682 | 0.144 | -0.433 | 0.665 |
| Middle and inferior temporal gyrus | -0.00927 | 0.144 | 1.47 | 0.142 |
| Hippocampus | -0.00137 | 0.144 | -0.474 | 0.635 |
| Insula | -0.0206 | 0.144 | -0.0640 | 0.949 |
| Nucleus accumbens | -0.0448 | 0.144 | -0.0100 | 0.992 |
| Cuneus | 0.202 | 0.144 | -0.143 | 0.886 |
| Lingual gyrus | 0.153 | 0.144 | -0.3^11^ | 0.756 |
| Lateral remainder of the occipital lobe | -0.100 | 0.144 | 1.40 | 0.161 |
| Pallidum | 0.0930 | 0.144 | 1.06 | 0.288 |
| Postcentral gyrus | 0.0380 | 0.144 | -0.695 | 0.487 |
| Inferiolateral remainder of parietal lobe | -0.0507 | 0.144 | 0.647 | 0.5^18^ |
| Superior parietal gyrus | 0.0690 | 0.144 | 0.264 | 0.792 |
| Posterior temporal lobe | 0.^18^5 | 0.144 | -0.352 | 0.725 |
| Presubgenual frontal cortex | -0.0284 | 0.144 | 0.480 | 0.631 |
| Putamen | -0.134 | 0.144 | 1.29 | 0.198 |
| Substantia nigra | 0.0531 | 0.144 | -0.197 | 0.843 |
| Subcallosal area | 0.163 | 0.144 | -0.931 | 0.352 |
| Subgenual frontal cortex | 0.00106 | 0.144 | 0.370 | 0.712 |
| Thalamus | -0.0362 | 0.00220 | 1.14 | 0.256 |
| Age | 0.001055 | 0.0323 | 0.480 | 0.631 |
| Sex | -0.03622 | 0.032336 | -1.12 | 0.263 |
| **Two factor interaction model (z-score ~ diagnostic group:tracer + region + age + sex)** | | | | |
| **Contrast** | **Estimate** | **Standard error** | **t ratio** | **p** |
| AD[^11^C]PK11195 – AD[^18^F]GE-180 | 0.0983 | 0.0477 | 2.06 | 0.308 |
| AD[^11^C]PK11195 – AD[^11^C]PBR28 | 0.222 | 0.0529 | 4.19 | 0.000400 |
| AD[^18^F]GE-180 – AD[^11^C]PBR28 | 0.123 | 0.0509 | 2.42 | 0.148 |
| Control[^11^C]PK11195 – Control[^18^F]GE-180 | -0.00108 | 0.0685 | -0.0160 | 1 |
| Control[^11^C]PK11195 – Control[^11^C]PBR28 | 0.00573 | 0.0651 | 0.0880 | 1 |
| Control[^18^F]GE-180 – Control[^11^C]PBR28 | 0.00682 | 0.0608 | 0.^11^2 | 1 |
| Precentral gyrus | ref | *ref* | ref | ref |
| Amygdala | 0.301 | 0.144 | 2.09 | 0.0366 |
| Anterior temporal lobe lateral part | -0.0682 | 0.144 | -0.474 | 0.635 |
| Anterior temporal lobe medial part | 0.0675 | 0.144 | 0.470 | 0.639 |
| Midbrain | 0.109 | 0.144 | 0.758 | 0.449 |
| Pons | 0.123 | 0.144 | 0.857 | 0.391 |
| Caudate nucleus | -0.381 | 0.144 | -2.65 | 0.00801 |
| Cerebellum dentate | 0.217 | 0.144 | 1.51 | 0.131 |
| Cerebellum white matter | 0.158 | 0.144 | 1.10 | 0.272 |
| Inferior frontal gyrus | 0.0166 | 0.144 | 0.^11^5 | 0.908 |
| Middle frontal gyrus | 0.132 | 0.144 | 0.922 | 0.357 |
| Anterior orbital gyrus | 0.141 | 0.144 | 0.984 | 0.325 |
| Lateral orbital gyrus | -0.00^11^7 | 0.144 | -0.00800 | 0.993 |
| Medial orbital gyrus | -0.0170 | 0.144 | -0.^118^ | 0.906 |
| Posterior orbital gyrus | -0.0316 | 0.144 | -0.220 | 0.826 |
| Straight gyrus | -0.0861 | 0.144 | -0.599 | 0.549 |
| Superior frontal gyrus | -0.0133 | 0.144 | -0.0930 | 0.926 |
| Cingulate gyrus anterior part | -0.129 | 0.144 | -0.897 | 0.370 |
| Cingulate gyrus posterior part | 0.0978 | 0.144 | 0.680 | 0.496 |
| Fusiform gyrus | 0.0882 | 0.144 | 0.614 | 0.539 |
| Parahippocampal and ambient gyri | 0.120 | 0.144 | 0.832 | 0.405 |
| Superior temporal gyrus anterior part | -0.316 | 0.144 | -2.20 | 0.0280 |
| Superior temporal gyrus posterior part | -0.0623 | 0.144 | -0.433 | 0.665 |
| Middle and inferior temporal gyrus | 0.2^11^ | 0.144 | 1.47 | 0.142 |
| Hippocampus | -0.0682 | 0.144 | -0.475 | 0.635 |
| Insula | -0.00927 | 0.144 | -0.0650 | 0.949 |
| Nucleus accumbens | -0.00137 | 0.144 | -0.0100 | 0.992 |
| Cuneus | -0.0206 | 0.144 | -0.143 | 0.886 |
| Lingual gyrus | -0.0448 | 0.144 | -0.312 | 0.755 |
| Lateral remainder of the occipital lobe | 0.202 | 0.144 | 1.40 | 0.161 |
| Pallidum | 0.153 | 0.144 | 1.06 | 0.288 |
| Postcentral gyrus | -0.100 | 0.144 | -0.696 | 0.487 |
| Inferiolateral remainder of parietal lobe | 0.0930 | 0.144 | 0.647 | 0.517 |
| Superior parietal gyrus | 0.0380 | 0.144 | 0.264 | 0.791 |
| Posterior temporal lobe | -0.0507 | 0.144 | -0.353 | 0.724 |
| Presubgenual frontal cortex | 0.0690 | 0.144 | 0.480 | 0.631 |
| Putamen | 0.^18^5 | 0.144 | 1.29 | 0.198 |
| Substantia nigra | -0.0284 | 0.144 | -0.198 | 0.843 |
| Subcallosal area | -0.134 | 0.144 | -0.931 | 0.352 |
| Subgenual frontal cortex | 0.0531 | 0.144 | 0.370 | 0.712 |
| Thalamus | 0.163 | 0.144 | 1.14 | 0.256 |
| Age | 0.000637 | 0.002204 | 0.289 | 0.773 |
| Sex | -0.0304 | 0.0328 | -0.929 | 0.353 |
| **Three factor interaction model (z-score ~ diagnostic group:tracer:region + age + sex)** | | | | |
| **Contrast** | **Estimate** | **Standard error** | **t ratio** | **p** |
| *Tracer: [^11^C]PK11195. Region: Precentral gyrus* | *ref* | *ref* | *ref* | *ref* |
| [^18^F]GE-180:Diagnostic group:Amygdala | 0.169 | 0.753 | 0.224 | 0.823 |
| [^11^C]PBR28:Diagnostic group:Amygdala | 1.50 | 0.758 | 1.98 | 0.0478 |
| [^18^F]GE-180:Diagnostic group:Anterior temporal lobe lateral part | 0.207 | 0.753 | 0.274 | 0.784 |
| [^11^C]PBR28:Diagnostic group:Anterior temporal lobe lateral part | 0.840 | 0.758 | 1.108 | 0.268 |
| [^18^F]GE-180:Diagnostic group:Anterior temporal lobe medial part | 0.662 | 0.753 | 0.88 | 0.379 |
| [^11^C]PBR28:Diagnostic group:Anterior temporal lobe medial part | 1.35 | 0.758 | 1.78 | 0.0744 |
| [^18^F]GE-180:Diagnostic group:Midbrain | 0.^11^6 | 0.753 | 0.155 | 0.877 |
| [^11^C]PBR28:Diagnostic group:Midbrain | 0.946 | 0.758 | 1.25 | 0.212 |
| [^18^F]GE-180:Diagnostic group:Pons | 0.830 | 0.753 | 1.103 | 0.270 |
| [^11^C]PBR28:Diagnostic group:Pons | 1.43 | 0.758 | 1.90 | 0.0590 |
| [^18^F]GE-180:Diagnostic group:Caudate nucleus | 0.437 | 0.753 | 0.581 | 0.561 |
| [^11^C]PBR28:Diagnostic group:Caudate nucleus | -0.190 | 0.758 | -0.251 | 0.802 |
| [^18^F]GE-180:Diagnostic group:Cerebellum dentate | 0.740 | 0.753 | 0.983 | 0.326 |
| [^11^C]PBR28:Diagnostic group:Cerebellum dentate | 1.14 | 0.758 | 1.5 | 0.134 |
| [^18^F]GE-180:Diagnostic group:Cerebellum white matter | 0.609 | 0.753 | 0.808 | 0.419 |
| [^11^C]PBR28:Diagnostic group:Cerebellum white matter | 0.652 | 0.758 | 0.86 | 0.390 |
| [^18^F]GE-180:Diagnostic group:Inferior frontal gyrus | -0.229 | 0.753 | -0.304 | 0.761 |
| [^11^C]PBR28:Diagnostic group:Inferior frontal gyrus | 0.439 | 0.758 | 0.579 | 0.563 |
| [^18^F]GE-180:Diagnostic group:Middle frontal gyrus | 0.0423 | 0.753 | 0.056 | 0.955 |
| [^11^C]PBR28:Diagnostic group:Middle frontal gyrus | 0.493 | 0.758 | 0.65 | 0.515 |
| [^18^F]GE-180:Diagnostic group:Anterior orbital gyrus | 0.599 | 0.753 | 0.796 | 0.426 |
| [^11^C]PBR28:Diagnostic group:Anterior orbital gyrus | 0.932 | 0.758 | 1.23 | 0.219 |
| [^18^F]GE-180:Diagnostic group:Lateral orbital gyrus | 0.224 | 0.753 | 0.297 | 0.767 |
| [^11^C]PBR28:Diagnostic group:Lateral orbital gyrus | 0.722 | 0.758 | 0.953 | 0.341 |
| [^18^F]GE-180:Diagnostic group:Medial orbital gyrus | 0.399 | 0.753 | 0.531 | 0.596 |
| [^11^C]PBR28:Diagnostic group:Medial orbital gyrus | 1.03 | 0.758 | 1.36 | 0.174 |
| [^18^F]GE-180:Diagnostic group:Posterior orbital gyrus | 0.505 | 0.753 | 0.67 | 0.503 |
| [^11^C]PBR28:Diagnostic group:Posterior orbital gyrus | 0.957 | 0.758 | 1.26 | 0.207 |
| [^18^F]GE-180:Diagnostic group:Straight gyrus | 0.100 | 0.753 | 0.133 | 0.894 |
| [^11^C]PBR28:Diagnostic group:Straight gyrus | 0.459 | 0.758 | 0.606 | 0.545 |
| [^18^F]GE-180:Diagnostic group:Superior frontal gyrus | -0.413 | 0.753 | -0.548 | 0.583 |
| [^11^C]PBR28:Diagnostic group:Superior frontal gyrus | 0.0236 | 0.758 | 0.031 | 0.975 |
| [^18^F]GE-180:Diagnostic group:Cingulate gyrus anterior part | -0.0322 | 0.753 | -0.043 | 0.966 |
| [^11^C]PBR28:Diagnostic group:Cingulate gyrus anterior part | 0.260 | 0.758 | 0.343 | 0.732 |
| [^18^F]GE-180:Diagnostic group:Cingulate gyrus posterior part | -0.2^18^ | 0.753 | -0.29 | 0.772 |
| [^11^C]PBR28:Diagnostic group:Cingulate gyrus posterior part | 0.370 | 0.758 | 0.488 | 0.625 |
| [^18^F]GE-180:Diagnostic group:Fusiform gyrus | 0.0265 | 0.753 | 0.035 | 0.972 |
| [^11^C]PBR28:Diagnostic group:Fusiform gyrus | 0.852 | 0.758 | 1.12 | 0.261 |
| [^18^F]GE-180:Diagnostic group:Parahippocampal and ambient gyri | 0.323 | 0.753 | 0.428 | 0.668 |
| [^11^C]PBR28:Diagnostic group:Parahippocampal and ambient gyri | 0.958 | 0.758 | 1.26 | 0.206 |
| [^18^F]GE-180:Diagnostic group:Superior temporal gyrus anterior part | -0.0758 | 0.753 | -0.101 | 0.920 |
| [^11^C]PBR28:Diagnostic group:Superior temporal gyrus anterior part | 0.975 | 0.758 | 1.29 | 0.198 |
| [^18^F]GE-180:Diagnostic group:Superior temporal gyrus posterior part | -0.334 | 0.753 | -0.444 | 0.657 |
| [^11^C]PBR28:Diagnostic group:Superior temporal gyrus posterior part | 0.385 | 0.758 | 0.508 | 0.612 |
| [^18^F]GE-180:Diagnostic group:Middle and inferior temporal gyrus | 0.376 | 0.753 | 0.5 | 0.617 |
| [^11^C]PBR28:Diagnostic group:Middle and inferior temporal gyrus | 0.840 | 0.758 | 1.109 | 0.268 |
| [^18^F]GE-180:Diagnostic group:Hippocampus | -0.990 | 0.753 | -1.31 | 0.^18^8 |
| [^11^C]PBR28:Diagnostic group:Hippocampus | 0.0695 | 0.758 | 0.092 | 0.927 |
| [^18^F]GE-180:Diagnostic group:Insula | -0.0383 | 0.753 | -0.051 | 0.959 |
| [^11^C]PBR28:Diagnostic group:Insula | 0.448 | 0.758 | 0.591 | 0.554 |
| [^18^F]GE-180:Diagnostic group:Nucleus accumbens | -0.476 | 0.753 | -0.633 | 0.527 |
| [^11^C]PBR28:Diagnostic group:Nucleus accumbens | 0.155 | 0.758 | 0.204 | 0.838 |
| [^18^F]GE-180:Diagnostic group:Cuneus | 0.168 | 0.753 | 0.223 | 0.824 |
| [^11^C]PBR28:Diagnostic group:Cuneus | -0.207 | 0.758 | -0.274 | 0.784 |
| [^18^F]GE-180:Diagnostic group:Lingual gyrus | 0.533 | 0.753 | 0.708 | 0.479 |
| [^11^C]PBR28:Diagnostic group:Lingual gyrus | 0.207 | 0.758 | 0.273 | 0.785 |
| [^18^F]GE-180:Diagnostic group:Lateral remainder of the occipital lobe | 0.459 | 0.753 | 0.609 | 0.542 |
| [^11^C]PBR28:Diagnostic group:Lateral remainder of the occipital lobe | 0.3^11^ | 0.758 | 0.41 | 0.682 |
| [^18^F]GE-180:Diagnostic group:Pallidum | 0.623 | 0.753 | 0.828 | 0.408 |
| [^11^C]PBR28:Diagnostic group:Pallidum | 0.907 | 0.758 | 1.20 | 0.232 |
| [^18^F]GE-180:Diagnostic group:Postcentral gyrus | -0.0892 | 0.753 | -0.^11^9 | 0.906 |
| [^11^C]PBR28:Diagnostic group:Postcentral gyrus | -0.00301 | 0.758 | -0.004 | 0.997 |
| [^18^F]GE-180:Diagnostic group:Inferiolateral remainder of parietal lobe | -0.237 | 0.753 | -0.314 | 0.753 |
| [^11^C]PBR28:Diagnostic group:Inferiolateral remainder of parietal lobe | -0.^18^4 | 0.758 | -0.243 | 0.808 |
| [^18^F]GE-180:Diagnostic group:Superior parietal gyrus | -0.0565 | 0.753 | -0.075 | 0.940 |
| [^11^C]PBR28:Diagnostic group:Superior parietal gyrus | 0.149 | 0.758 | 0.197 | 0.844 |
| [^18^F]GE-180:Diagnostic group:Posterior temporal lobe | 0.194 | 0.753 | 0.257 | 0.797 |
| [^11^C]PBR28:Diagnostic group:Posterior temporal lobe | 0.^11^2 | 0.758 | 0.148 | 0.882 |
| [^18^F]GE-180:Diagnostic group:Presubgenual frontal cortex | 0.0898 | 0.753 | 0.^11^9 | 0.905 |
| [^11^C]PBR28:Diagnostic group:Presubgenual frontal cortex | 0.198 | 0.758 | 0.261 | 0.794 |
| [^18^F]GE-180:Diagnostic group:Putamen | 0.444 | 0.753 | 0.589 | 0.556 |
| [^11^C]PBR28:Diagnostic group:Putamen | 0.787 | 0.758 | 1.04 | 0.299 |
| [^18^F]GE-180:Diagnostic group:Substantia nigra | 0.0803 | 0.753 | 0.107 | 0.915 |
| [^11^C]PBR28:Diagnostic group:Substantia nigra | 0.586 | 0.758 | 0.772 | 0.440 |
| [^18^F]GE-180:Diagnostic group:Subcallosal area | -0.896 | 0.753 | -1.19 | 0.234 |
| [^11^C]PBR28:Diagnostic group:Subcallosal area | -0.0929 | 0.758 | -0.123 | 0.903 |
| [^18^F]GE-180:Diagnostic group:Subgenual frontal cortex | 0.0362 | 0.753 | 0.048 | 0.962 |
| [^11^C]PBR28:Diagnostic group:Subgenual frontal cortex | -0.127 | 0.758 | -0.167 | 0.867 |
| [^18^F]GE-180:Diagnostic group:Thalamus | 0.379 | 0.753 | 0.503 | 0.615 |
| [^11^C]PBR28:Diagnostic group:Thalamus | 1.01 | 0.758 | 1.34 | 0.^18^2 |
| *Tracer: [^11^C]GE-180. Region: Precentral gyrus* | ref | *ref* | *ref* | ref |
| [^11^C]PBR28:Diagnostic group:Amygdala | 1.33 | 0.713 | 1.87 | 0.0619 |
| [^11^C]PBR28:Diagnostic group:Anterior temporal lobe lateral part | 0.633 | 0.713 | 0.888 | 0.375 |
| [^11^C]PBR28:Diagnostic group:Anterior temporal lobe medial part | 0.690 | 0.713 | 0.967 | 0.333 |
| [^11^C]PBR28:Diagnostic group:Midbrain | 0.830 | 0.713 | 1.16 | 0.245 |
| [^11^C]PBR28:Diagnostic group:Pons | 0.601 | 0.713 | 0.843 | 0.399 |
| [^11^C]PBR28:Diagnostic group:Caudate nucleus | -0.628 | 0.713 | -0.88 | 0.379 |
| [^11^C]PBR28:Diagnostic group:Cerebellum dentate | 0.398 | 0.713 | 0.557 | 0.577 |
| [^11^C]PBR28:Diagnostic group:Cerebellum white matter | 0.0436 | 0.713 | 0.061 | 0.951 |
| [^11^C]PBR28:Diagnostic group:Inferior frontal gyrus | 0.667 | 0.713 | 0.935 | 0.350 |
| [^11^C]PBR28:Diagnostic group:Middle frontal gyrus | 0.451 | 0.713 | 0.632 | 0.528 |
| [^11^C]PBR28:Diagnostic group:Anterior orbital gyrus | 0.333 | 0.713 | 0.466 | 0.641 |
| [^11^C]PBR28:Diagnostic group:Lateral orbital gyrus | 0.499 | 0.713 | 0.699 | 0.484 |
| [^11^C]PBR28:Diagnostic group:Medial orbital gyrus | 0.630 | 0.713 | 0.884 | 0.377 |
| [^11^C]PBR28:Diagnostic group:Posterior orbital gyrus | 0.452 | 0.713 | 0.634 | 0.526 |
| [^11^C]PBR28:Diagnostic group:Straight gyrus | 0.359 | 0.713 | 0.503 | 0.615 |
| [^11^C]PBR28:Diagnostic group:Superior frontal gyrus | 0.437 | 0.713 | 0.612 | 0.541 |
| [^11^C]PBR28:Diagnostic group:Cingulate gyrus anterior part | 0.292 | 0.713 | 0.409 | 0.682 |
| [^11^C]PBR28:Diagnostic group:Cingulate gyrus posterior part | 0.589 | 0.713 | 0.825 | 0.410 |
| [^11^C]PBR28:Diagnostic group:Fusiform gyrus | 0.825 | 0.713 | 1.16 | 0.247 |
| [^11^C]PBR28:Diagnostic group:Parahippocampal and ambient gyri | 0.636 | 0.713 | 0.891 | 0.373 |
| [^11^C]PBR28:Diagnostic group:Superior temporal gyrus anterior part | 1.05 | 0.713 | 1.47 | 0.141 |
| [^11^C]PBR28:Diagnostic group:Superior temporal gyrus posterior part | 0.719 | 0.713 | 1.01 | 0.314 |
| [^11^C]PBR28:Diagnostic group:Middle and inferior temporal gyrus | 0.464 | 0.713 | 0.65 | 0.516 |
| [^11^C]PBR28:Diagnostic group:Hippocampus | 1.06 | 0.713 | 1.49 | 0.137 |
| [^11^C]PBR28:Diagnostic group:Insula | 0.487 | 0.713 | 0.682 | 0.495 |
| [^11^C]PBR28:Diagnostic group:Nucleus accumbens | 0.631 | 0.713 | 0.885 | 0.376 |
| [^11^C]PBR28:Diagnostic group:Cuneus | -0.375 | 0.713 | -0.526 | 0.599 |
| [^11^C]PBR28:Diagnostic group:Lingual gyrus | -0.326 | 0.713 | -0.457 | 0.648 |
| [^11^C]PBR28:Diagnostic group:Lateral remainder of the occipital lobe | -0.148 | 0.713 | -0.207 | 0.836 |
| [^11^C]PBR28:Diagnostic group:Pallidum | 0.283 | 0.713 | 0.397 | 0.691 |
| [^11^C]PBR28:Diagnostic group:Postcentral gyrus | 0.0862 | 0.713 | 0.121 | 0.904 |
| [^11^C]PBR28:Diagnostic group:Inferiolateral remainder of parietal lobe | 0.0523 | 0.713 | 0.073 | 0.942 |
| [^11^C]PBR28:Diagnostic group:Superior parietal gyrus | 0.206 | 0.713 | 0.289 | 0.773 |
| [^11^C]PBR28:Diagnostic group:Posterior temporal lobe | -0.0815 | 0.713 | -0.^11^4 | 0.909 |
| [^11^C]PBR28:Diagnostic group:Presubgenual frontal cortex | 0.108 | 0.713 | 0.151 | 0.880 |
| [^11^C]PBR28:Diagnostic group:Putamen | 0.343 | 0.713 | 0.481 | 0.630 |
| [^11^C]PBR28:Diagnostic group:Substantia nigra | 0.505 | 0.713 | 0.708 | 0.479 |
| [^11^C]PBR28:Diagnostic group:Subcallosal area | 0.803 | 0.713 | 1.13 | 0.261 |
| [^11^C]PBR28:Diagnostic group:Subgenual frontal cortex | -0.163 | 0.713 | -0.228 | 0.820 |
| [^11^C]PBR28:Diagnostic group:Thalamus | 0.633 | 0.713 | 0.887 | 0.375 |
